# Genetic and non-genetic factors affecting the expression of COVID-19 relevant genes in the large airway epithelium

**DOI:** 10.1101/2020.10.01.20202820

**Authors:** Silva Kasela, Victor E Ortega, Molly Martorella, Suresh Garudadri, Jenna Nguyen, Elizabeth Ampleford, Anu Pasanen, Srilaxmi Nerella, Kristina L Buschur, Igor Z Barjaktarevic, R Graham Barr, Eugene R Bleecker, Russell P Bowler, Alejandro P Comellas, Christopher B Cooper, David J Couper, Gerard J Criner, Jeffrey L Curtis, MeiLan K Han, Nadia N Hansel, Eric A Hoffman, Robert J Kaner, Jerry A Krishnan, Fernando J Martinez, Merry-Lynn N McDonald, Deborah A Meyers, Robert Paine, Stephen P Peters, Mario Castro, Loren C Denlinger, Serpil C Erzurum, John V Fahy, Elliot Israel, Nizar N Jarjour, Bruce D Levy, Xingnan Li, Wendy C Moore, Sally E Wenzel, Joe Zein, NHLBI SubPopulations and InteRmediate Outcome Measures In COPD Study (SPIROMICS), NHLBI Trans-Omics for Precision Medicine (TOPMed) Consortium, Charles Langelier, Prescott G Woodruff, Tuuli Lappalainen, Stephanie A Christenson

## Abstract

Particular host and environmental factors influence susceptibility to severe COVID-19. We analyzed RNA-sequencing data from bronchial epithelial brushings - a relevant tissue for SARS-CoV-2 infection - obtained from three cohorts of uninfected individuals, and investigated how non-genetic and genetic factors affect the regulation of host genes implicated in COVID-19. We found that *ACE2* expression was higher in relation to active smoking, obesity, and hypertension that are known risk factors of COVID-19 severity, while an association with interferon-related inflammation was driven by the truncated, non-binding *ACE2* isoform. We discovered that expression patterns of a suppressed airway immune response to early SARS-CoV-2 infection, compared to other viruses, are similar to patterns associated with obesity, hypertension, and cardiovascular disease, which may thus contribute to a COVID-19-susceptible airway environment. eQTL mapping identified regulatory variants for genes implicated in COVID-19, some of which had pheWAS evidence for their potential role in respiratory infections. These data provide evidence that clinically relevant variation in the expression of COVID-19-related genes is associated with host factors, environmental exposures, and likely host genetic variation.

## Main text

Coronavirus disease 2019 (COVID-19), the clinical syndrome caused by the severe acute respiratory syndrome coronavirus 2 (SARS-CoV-2) virus, has led to a global crisis. As a respiratory virus, SARS-CoV-2 is hypothesized to gain entry into humans via the airway epithelium, where it initiates a host response that leads to the subsequent clinical syndrome. Despite an immense global burden of disease, the manifestations of SARS-CoV-2 infection vary enormously, from asymptomatic infection to progressive acute respiratory failure and death. The viral or host features that determine the course of disease in each individual are poorly understood. Multiple clinical risk factors for severe COVID-19 have been identified, including older age, male sex, African American race, smoking, and comorbidities such as hypertension, obesity, diabetes, cardiovascular disease, and chronic airway diseases^1–6^, as well as host genetics^6–9^.

The expression levels of genes that interact with the SARS-CoV-2 virus or are involved in the subsequent host response are hypothesized to be an important host factor that could partially underlie the substantial inter-individual variability in COVID-19 susceptibility and progression^10–12^. In this study, we analyzed genetic and non-genetic factors influencing the expression of human genes that have been implicated in COVID-19 (study design in **Extended Data Fig. 1**). We analyzed RNA-sequencing (RNA-seq) data from bronchial brushing samples obtained from the SPIROMICS cohort^13^ (*n* = 163), notable for the high burden of COVID-19-relevant comorbidities and rich phenotype and whole genome sequencing (WGS) data from the TOPMed Project^14^. For replication, we used two asthma RNA-seq datasets, SARP (*n* = 156) and MAST (*n* = 35) as well as eQTL data from GTEx^15^.

We first analyzed expression levels of *ACE2*, the receptor of the SARS-CoV-2 Spike protein that is the key host gene for viral entry^16,17^. Corroborating previous reports^12,18–20^, we found that current smoking, when compared to non-smoking, had the largest overall effect on *ACE2* expression of any phenotypic feature studied in SPIROMICS, before and after adjustments for covariates (log_2_ fold change (FC) = 0.30 ± 0.06, *P* = 1.7×10^-7^, **Fig. 1a, Supplementary Table 1**). This effect was absent in former smokers. In similarly adjusted models, we found no association between *ACE2* levels and Chronic Obstructive Pulmonary Disease (COPD, **Extended Data Fig. 2a**), nor with asthma in MAST^20^ (**Extended Data Fig. 2b**). In SARP, *ACE2* levels were slightly lower in asthmatics compared to healthy controls (**Extended Data Fig. 2c**), which was largely driven by decreased expression of *ACE2* only in asthmatics on oral steroids (**Extended Data Fig. 2d)**. African American race was associated with increased *ACE2* expression in both SPIROMICS and SARP, but no association after adjusting for covariates suggests that this was due to a higher prevalence of comorbid conditions (**Extended Data Fig. 2e-f**). However, *ACE2* expression was significantly higher across datasets in association with two relevant comorbidities, obesity and hypertension (**Fig. 1b-c, Extended Data Fig. 3a-e, Extended Data Fig. 4a-b**). Of note, we further found that use of anti-hypertensives in SPIROMICS attenuates the association between *ACE2* and hypertension towards levels seen in non-hypertensive participants (**Fig. 1c)**. When stratified by anti-hypertensive class, angiotensin receptor blockers (ARBs) and diuretics, but not ACE inhibitors or calcium channel blockers, were associated with lower *ACE2* levels, partially dependent on smoking status (**Extended Data Fig. 4c)**. Counterintuitively, modest decreases in *ACE2* expression were seen in SPIROMICS in association with age (log_2_ FC = -0.064 ± 0.02, *P* = 0.005 for every 10-year age increase, **Fig. 1e**) and male sex (log_2_ FC = -0.076 ± 0.035, *P* = 0.03, **Fig. 1d**) before and after adjustments, although similar associations were not seen in SARP or MAST.

**Fig. 1:**
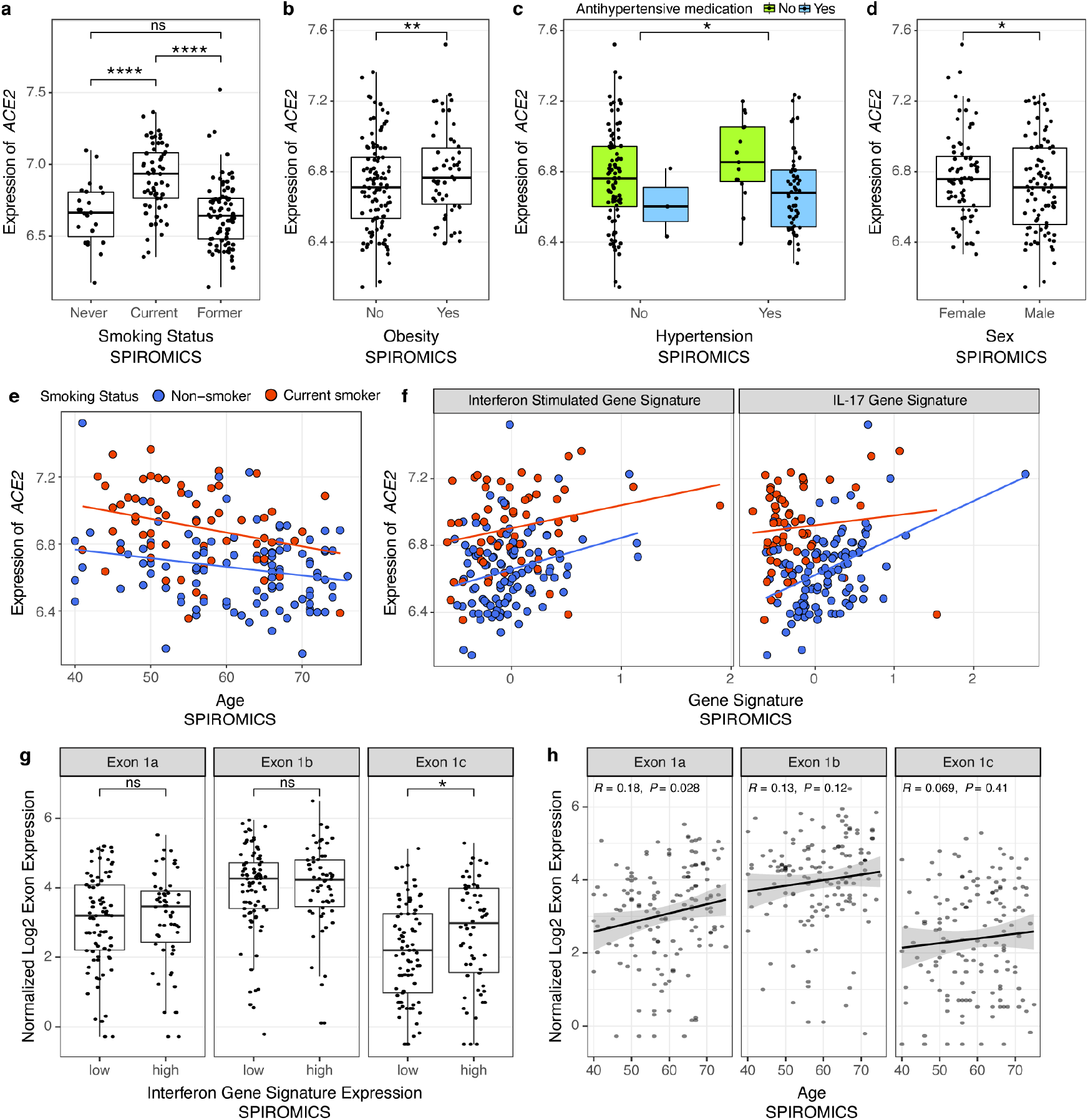
*ACE2* gene expression associations in SPIROMICS. **a-d**, Boxplots showing that *ACE2* log_2_ gene expression (*x*-axis) was increased in association with current but not former smoking as compared to never smokers (**a**), obesity (**b**, validated in the MAST and SARP cohorts, **Extended Data Fig. 3a-b**), hypertension (**c**, adjustments include anti-hypertensive treatment, validated in SARP, **Extended Data Fig. 4a**, data not collected in MAST), and female sex (**d**, not replicated in either MAST or SARP, data not shown). **e**, In SPIROMICS, *ACE2* gene expression was modestly decreased in association with advancing age, but this finding was not replicated in either MAST or SARP (**Supplementary Table 1A**). **f**, Scatterplots showing that *ACE2* gene expression increased in association with higher levels of our previously validated gene signatures of the airway epithelial response to interferon (left panel) and to IL-17 inflammation (right panel) after adjusting for smoking status. Both these findings were replicated in MAST and SARP (**Supplementary Table 1B**). **g**, Boxplots showing that ACE2 Exon 1c, which contributes to the truncated ACE2 transcript is differentially increased in association with our interferon signature while Exons 1a and 1b that contribute to the full length transcript are not associated. **h**, Scatterplots showing increased Exon 1a usage with advancing age that is not observed for Exon 1b and 1c. *P*-values indicated by: ****<0.0001, ***<0.001, **<0.01, *<0.05, ns=not significant in linear models adjusted for covariates.

As chronic airway inflammation, prevalent but heterogeneous in the airway diseases studied in the included cohorts, can influence gene expression and the host response to infections, we next studied how stereotypic adaptive airway immune responses affect *ACE2* expression. We used our previously validated gene expression signatures to quantify Type 2-, Interferon-, and IL-17-associated inflammation^21–23^. We found that *ACE2* expression was associated with increased interferon-related inflammation, as previously reported^10^, as well as IL-17-related but not Type 2 inflammation across datasets (**Fig. 1f**). Corroborating the association with IL-17 inflammation, genes highly co-expressed with *ACE2* expression included genes in our IL-17 signature across datasets (**Supplementary Table 2**). Recent reports suggested that *ACE2* induction by interferon stimulation may be explained by expression of a truncated *ACE2* isoform (initiated from exon 1c instead of 1a/b) that does not bind the SARS-CoV-2 spike protein^24,25^. We first corroborated this finding, showing that our interferon-stimulated gene signature is associated with increased exon 1c but not exons 1a or 1b usage (**Fig. 1g**). We also identified an increase in exon 1a usage with age suggesting that despite a decrease in overall ACE2 expression in association with age, expression of the full length ACE2 transcript may not be decreased (**Fig. 1h**). Importantly, differential exon 1c usage was not associated with any other clinical/biological outcomes of interest.

These results overall indicate that smoking, obesity, and hypertension affect airway epithelial expression of functional *ACE2* isoforms, as previously shown for smoking^12,18,19^. The *ACE2* association with interferon-related inflammation appears to be explained by the truncated version of *ACE2*^24,25^. Together these findings suggest that smoking, obesity and hypertension may contribute to COVID-19 severity through an association with increased *ACE2* expression, while other risk factors such as male sex and airway disease likely contribute via other mechanisms, corroborating recent evidence on sex differences in the immune response to COVID-19^26^.

As the host’s ability to mount an appropriate response to respiratory viruses may alter susceptibility to severe infection, we next performed gene set enrichment analyses (GSEA) to determine whether clinical risk factors are associated with similar airway gene expression patterns indicative of a diminished immune response that we recently identified early in COVID-19 by nasal/oropharyngeal swab^27^. As we previously reported^27^, the genes differentially expressed in association with SARS-CoV-2 infection compared to other viruses at diagnosis indicate a diminished innate and adaptive immune response that may allow for unabated viral infection and account for the long pre-symptomatic period associated with COVID-19. We hypothesized that clinical risk factors uniquely associated with COVID-19 severity (e.g. cardiovascular disease, hypertension) could predispose patients to develop more severe disease by contributing to this relative immunosuppression. We derived gene sets from our previously published RNA-seq data collected by nasal/oropharyngeal swab from patients at diagnosis of acute respiratory illness; 94 had COVID-19, 41 had other viral illness, and 103 had no virus identified by metagenomic sequencing analysis^27^. First, we generated gene sets derived from the 100 genes most up- and downregulated in association with infection type to use to determine if there were global similarities in gene expression changes across datasets. For pathway analyses, we then generated COVID-19 relevant gene sets specific to particular canonical pathways by inputting significantly differentially expressed genes (*FDR* < 0.05) between SARS-CoV-2 infection and other viral respiratory illness into the Ingenuity Pathway Analysis canonical pathway function (**Supplementary Table 3**). GSEA was then performed using FGSEA^28^ in which these gene sets were tested against gene lists ranked by their log fold change differential expression in association with comorbid clinical risk factors.

We found that the genes most downregulated in association with SARS-CoV-2 infection as compared to other viruses were significantly enriched amongst genes downregulated in association with obesity, hypertension, and cardiovascular disease in SPIROMICS (**Fig. 2a-c**). Findings for obesity were replicated in SARP and MAST, and for hypertension in SARP (**Extended Data Fig. 5a-c**, hypertension data not collected in MAST, cardiovascular disease data not collected in SARP or MAST). Conversely, genes upregulated in other viral infections (or conversely, downregulated by SARS-CoV-2) were upregulated in inflammatory airway conditions (current and former smokers, COPD) (**Fig. 2d-f**). Aging was associated with an enrichment in genes downregulated by SARS-CoV-2 infection only in MAST while genes upregulated with SARS-CoV-2 infection were enriched with increasing age across the datasets (**Extended Data Fig. 5d-f**). Our results demonstrate a sharp contrast between SARS-CoV-2 and other viral infections, which often trigger airway disease exacerbations by potentiating the chronic airway inflammation associated with these diseases and smoking exposure. We found this same pattern in association with asthma in MAST but not when considering asthma overall in SARP, potentially due to heterogeneity of its asthma subjects. When considering just asthmatics with uncontrolled symptoms or those on inhaled compared to no steroids (a marker of severity) we did find this same enrichment of genes up and downregulated in association with non-COVID viral infections (pathway enrichment shown in **Fig. 2g**).

**Fig. 2:**
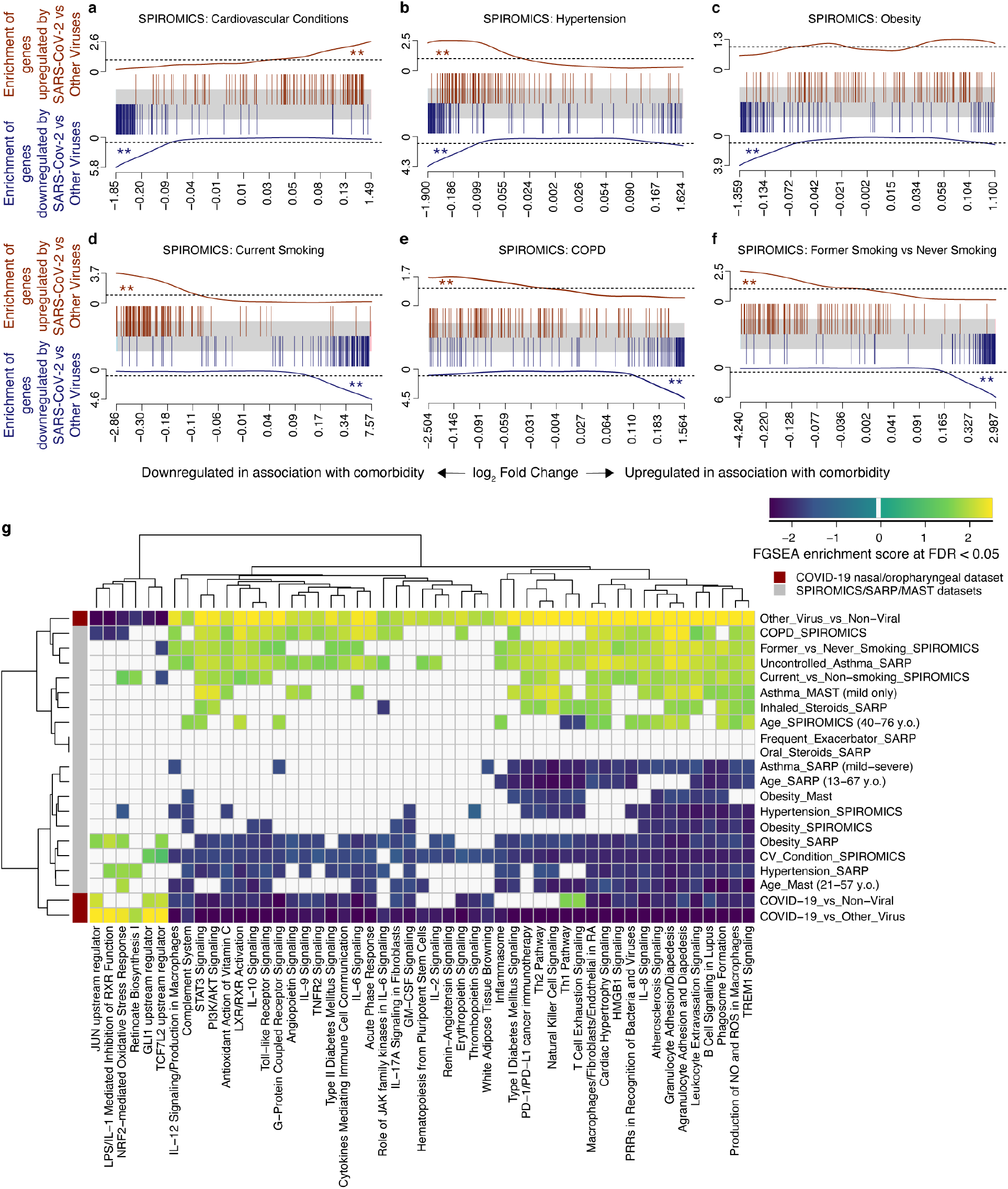
COVID-19-related gene set enrichment analyses in association with comorbidities. **a-f**, Barcode plots in which the vertical lines represent the 100 genes most upregulated (red) or downregulated (blue) in nasal/oropharyngeal swab samples obtained from COVID-19 patients as compared to other viruses at the time of diagnosis of an acute upper respiratory infection. These gene sets are plotted against log fold gene expression changes arranged from most downregulated to most upregulated with that comorbidity (horizontal grey bar). Lines above (red) and below (blue) the bar represent the running sum statistic with a significant finding indicated when the line crosses the dashed line at either end of the plot. Genes downregulated by SARS-CoV-2 infection compared to other viruses were significantly enriched amongst genes downregulated in association with cardiovascular conditions overall (**a**), hypertension (**b**), and obesity (**c**), while in Current (**d**) and Former smoking (**f**) and in COPD (**e**) these downregulated genes in COVID-19 were enriched among upregulated genes in association with comorbidity. ** indicates FDR < 0.05. **g**, COVID-19-related pathway gene sets were generated from an IPA analysis of the genes up and downregulated by SARS-CoV-2 infection compared to other viruses. Gene set enrichment scores for gene sets enriched at FDR < 0.05 (columns) are shown in the heatmap plotted against comorbidities (rows) with gene sets enriched amongst downregulated and upregulated genes indicated in blue and yellow, respectively. All pathways not enriched at FDR < 0.05 were shrunk to zero (white). Euclidean distance with average linkage was used for clustering.

We used pathway gene set enrichment to determine the potential biological significance of these findings. We found across datasets that pathway gene sets derived from genes downregulated by SARS-CoV-2 infection as compared to other viruses were also enriched amongst genes downregulated in association with obesity, hypertension, cardiovascular disease, and aging (FDR < 0.05, **Fig. 2g, Supplementary Table 4**). Enriched downregulated pathways included those related to pro-inflammatory cytokines such as IL-6 and IL-17 as well as macrophage and granulocyte activation. Furthermore, pathways related to cardiovascular and metabolic disease signaling such as atherosclerosis and diabetes signaling were also enriched. We confirmed the enriched findings by separately performing IPA canonical pathway analyses on the genes differentially expressed (*P* < 0.05) in association with these comorbidities, finding similar results in these global/unsupervised analyses (**Supplementary Table 5**). Conversely, pro-inflammatory airway conditions such as smoking and COPD led to opposite effects. These findings suggest that obesity, hypertension, cardiovascular disease, and age are associated with a relative COVID-19-relevant immunosuppression at the airway epithelium, which, by stunting early anti-viral host responses, could contribute to increased susceptibility to SARS-CoV-2 infection and disease severity.

In order to map host genetic variants, we focused on 496 genes implicated in SARS-CoV-2 infection (**Supplementary Table 6)**: *ACE2* and *TMPRSS2*, key genes for viral entry^16^; *CTSL, CTSB*, and *BSG*, which may have a role as alternative routes for viral entry^16,29^; host genes with protein-protein interactions with viral proteins^30^; differentially expressed genes as a response to the infection in cultured airway epithelial cells^31^; genes involved in autophagy that might counteract viral infection^32^; and other high interest genes from the COVID-19 Cell Atlas (*www.covid19cellatlas.org*). Our *cis*-eQTL mapping in SPIROMICS (*n* = 144) identified significant (genome-wide FDR < 0.05) genetic regulatory variation for 108 (21.8%) of these COVID-19-related genes (**Fig. 3a, Supplementary Table 7**), with many genes also having significant eQTLs in other tissues in GTEx^15^ (**Supplementary Table 8**). Given the sample size, we have good power to discover the vast majority of eQTLs with >2-fold effect on gene expression^15^. Many of the genes have a substantial genetic effect on gene expression: for example, the MERS receptor *DPP4*^33^ has a *cis*-regulatory variant rs6727102 where the alternative allele decreases expression by 3.3-fold (**Fig. 3a**). In 16 genes, the genetic regulatory effects were >50% of the magnitude of the differential expression induced by SARS-CoV-2 infection (**Fig. 3b**). While the key genes *ACE2* or *TMPRSS2* did not have eQTLs in bronchial epithelium (**Extended Data Fig. 6a-b**), as previously reported^20^, *TMPRSS2* has an eQTL in GTEx lung tissue. This is consistent with the lack of phenome-wide association signals^34^ or COVID-19 GWAS association at these loci^9^, suggesting that genetic regulation of these two genes is unlikely to contribute to potential host genetic effects on COVID-19. Many of the genes analyzed for eQTLs had variation in expression associated to clinical factors and comorbidities, with current smoking associated with the highest number of up- and down-regulated genes in association with comorbidity (**Extended Data Fig. 7a-b**). Compared to *ACE2*, the effect of current smoking on the expression of *TMPRSS2* was modest (**Extended Data Fig. 6c**), and as previously reported^11^, expression levels of *TMPRSS2* were higher in asthmatic than healthy controls, but not in COPD, and it decreased in association with steroid use (**Extended Data Fig. 6d**).

**Fig. 3:**
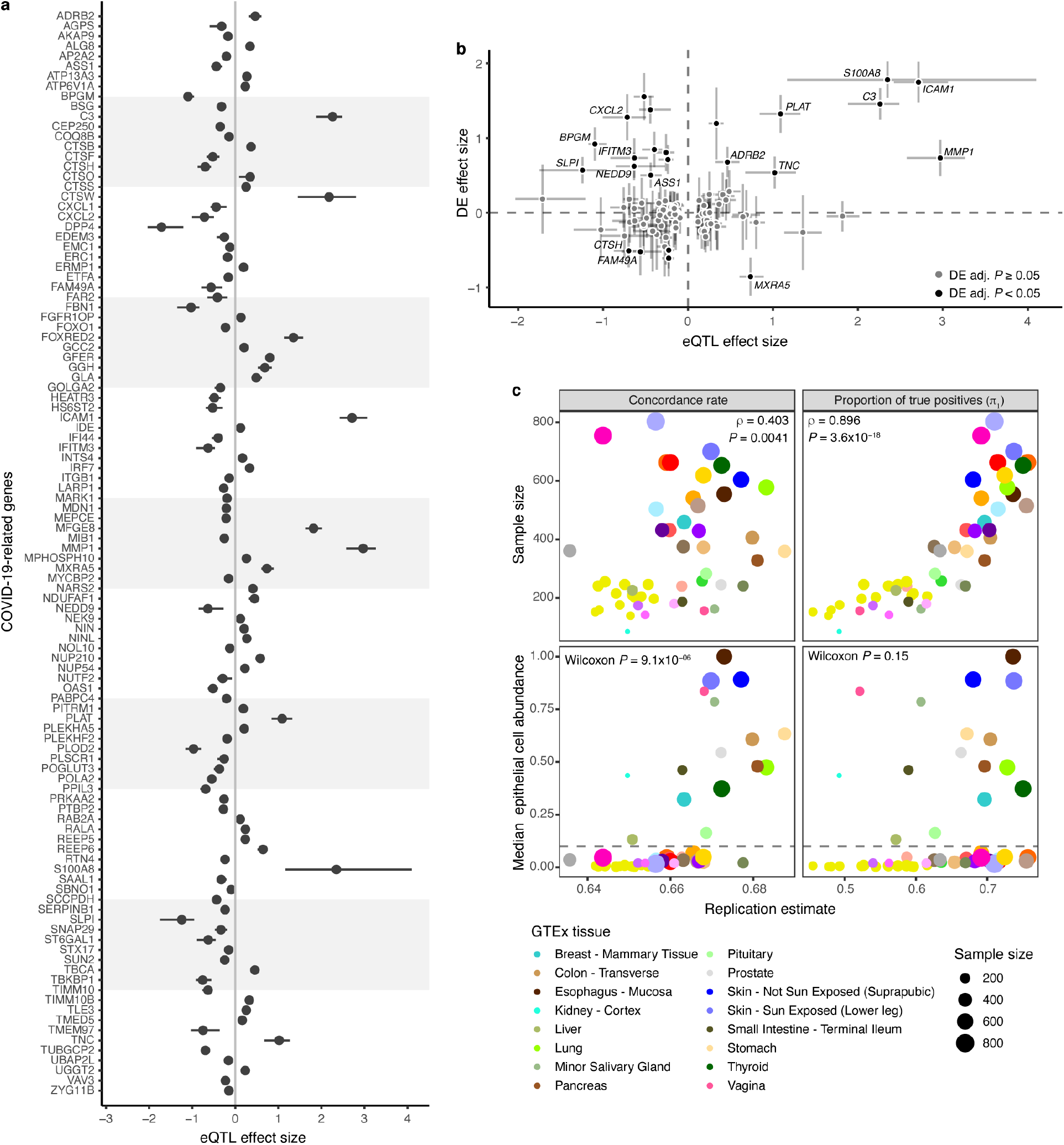
eQTLs in bronchial epithelium. **a**, Effect size measured as allelic fold change (aFC, log_2_) of the significant eQTLs for COVID-19 candidate genes. Error bars denote 95% bootstrap confidence intervals. **b**, Comparison of the regulatory effects and the effect of SARS-CoV-2 infection on the transcription of COVID-19 candidate genes in normal bronchial epithelial cells from Blanco-Melo et al.^31^ The graph shows regulatory effects as aFC as in **a**, and fold change (log_2_) of differential expression comparing the infected with mock-treated cells with error bars denoting the 95% confidence interval. Genes with adjusted *P*-value < 0.05 in the differential expression analysis are colored in black, genes with non-significant effect are colored in grey. Highlighted genes have eQTL effect size greater than 50% of the differential expression effect size on the absolute scale. DE - differential expression. **c**, Replication of *cis*-eQTLs from bronchial epithelium in GTEx v8 using the concordance rate (proportion of gene-variant pairs with the same direction of the effect, left panel) and proportion of true positives (*π*_1_, right panel). Upper panel shows the effect of sample size on the replication and concordance measures quantified as Spearman correlation coefficient (ρ). Lower panel shows the replication and concordance measures as the function of epithelial cell enrichment of the tissues measured as median epithelial cell enrichment score from xCell. Grey dashed line denotes median enrichment score > 0.1, which classifies tissues as enriched for epithelial cells. Wilcoxon rank sum test was used to estimate the difference in replication estimates between tissues enriched or not enriched for epithelial cells. The 16 tissues enriched for epithelial cells are outlined in the figure legend, for the full color legend see **Extended Data Fig. 8**.

*Cis*-eQTLs from bronchial epithelium replicated at a high rate in those tissues from the GTEx v8 dataset^15^ that have a large sample size or high epithelial cell abundance (**Fig. 3c, Extended Data Fig. 8**), reflecting similarity in cell type composition manifesting in similarity of regulatory variant activity^15^. However, relative to GTEx lung, our bronchial epithelium eQTLs included genes enriched for sensory perception of chemical stimulus and smell (**Supplementary Table 9**). In total of 143 genes with eQTLs in SPIROMICS were not tested in GTEx nor eQTLGen Consortium^35^, since bronchial epithelium is not well represented in previous eQTL catalogs. In addition to standard *cis*-eQTL mapping, we mapped cell type interacting eQTLs^36^ but none were discovered for the COVID-19-related genes.

To study the role of these regulatory variants in COVID-19 risk, we first analyzed eQTLs in the chromosome 3 locus with a significant association with hospitalization due to COVID-19^9^ and severe COVID-19 with respiratory failure^6,8^. We found no significant eQTLs in the bronchial epithelium for any of the six genes in this locus (**Extended Data Fig 6e**), suggesting that this genetic association may be driven by other tissues or cell types with a role in COVID-19. Next, given that COVID-19 GWAS still have limited power, we analyzed how regulatory variants for COVID-19 relevant genes associate to other immune- or respiratory-related phenotypes in large GWAS. Indication of these variants affecting (respiratory) infections would provide hypotheses of variants that might play a role in COVID-19 risk and its comorbidities (**Fig. 4a**). Thus, we performed a pheWAS analysis by Phenoscanner v2^37,38^ for the 108 lead *cis*-eQTLs for COVID19-related genes and diverse set of phenotypes (**Supplementary Table 10)**. Furthermore, we used the SPIROMICS phenotype data to study associations for 20 phenotypes (**Supplementary Table 11**). Of these loci, 44 were associated with at least one phenotype (*P* < 10^-5^), with expected patterns - best powered GWAS traits having most associations and shared signals for highly correlated traits (**Extended Data Fig. 9**). We further used colocalization analysis to extract loci where the eQTL and GWAS signals are likely to share a causal variant, as opposed to spurious overlap, focusing on 20 loci with associations for hematological and respiratory system traits of which 12 colocalized (PP4 > 0.5, **Fig. 4b, Supplementary Table 12**). In **Figure 4c**, we show Interferon-induced transmembrane protein 3 (*IFITM3)* where the eQTL is associated with multiple blood cell traits of the immune system^39^ and neutrophil counts in SPIROMICS (*P* < 0.002). This gene is upregulated by SARS-CoV-2 infection^31^ and has a well-characterized role in the entry of multiple viruses, including coronaviruses^40^. Another interesting gene, *ERMP1* (**Fig. 4d**) has an eQTL colocalizing with an asthma GWAS association in the UK Biobank. ERMP1 interacts with the SARS-CoV-2 protein Orf9c^30^, and severe asthma is a risk factor for COVID-19 hospitalization^6^ and death^41^. An eQTL for the *MEPCE* gene that interacts with SARS-Cov-2 protein Nsp8^30^ is associated with platelet parameters^39^ (**Fig. 4e**). Interestingly, platelets are hyperactivated in COVID-19^42,43^ and platelet count could be used as a prognostic biomarker in COVID-19 patients^44–46^.

**Fig. 4:**
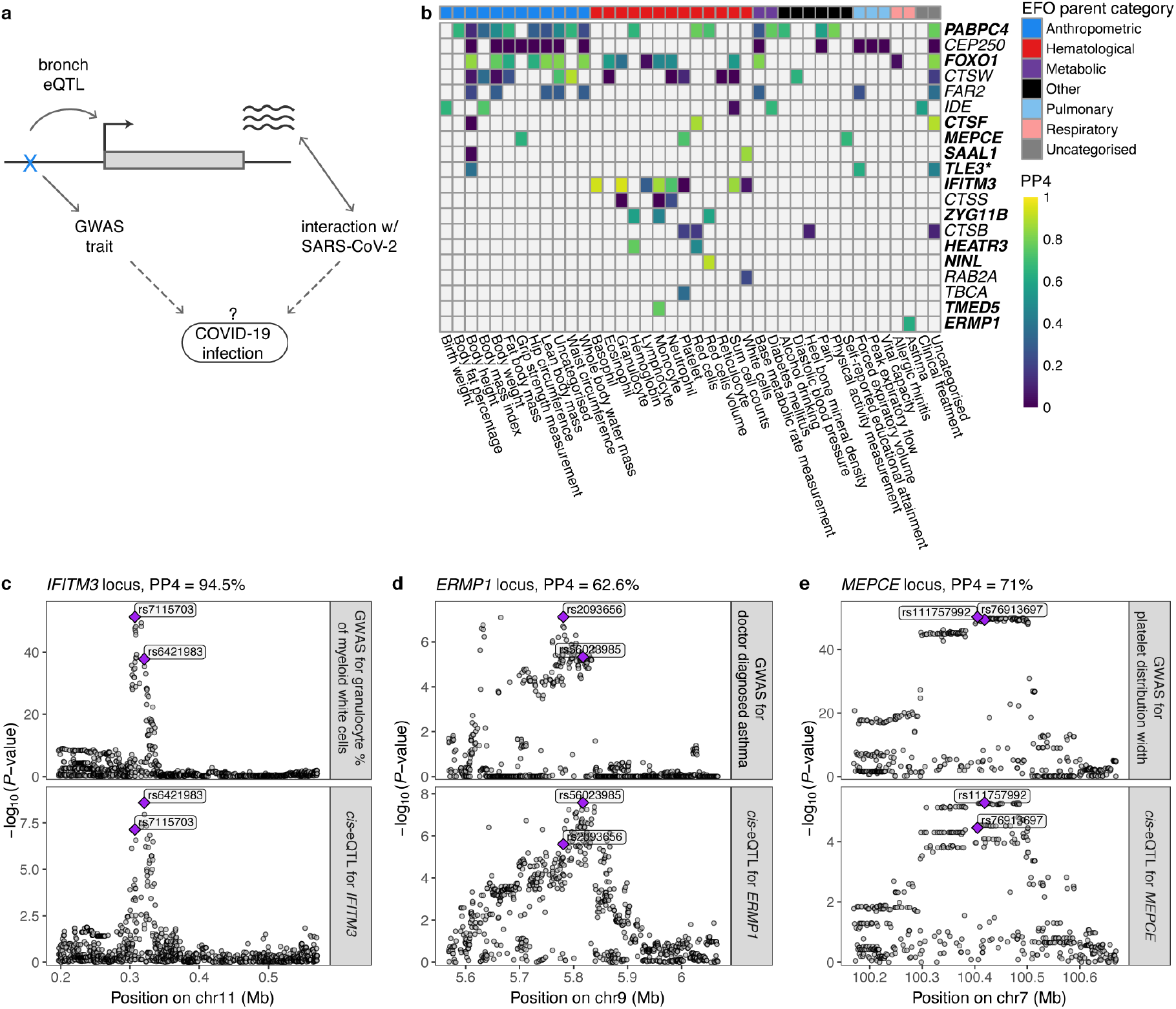
Colocalization analysis of the regulatory variants for COVID-19-related genes. **a**, Illustration of the concept of how regulatory variants for COVID-19-related genes in bronchial epithelium can be possible candidates for genetic factors that affect infection or progression of the disease. Dotted lines denote the hypothesis we are able to create by searching for the phenotypic associations of the eQTLs for COVID-19-related genes. **b**, Heatmap of the colocalization analysis results for 20 COVID-19-related genes with eQTLs that have at least one phenotypic association belonging to the experimental factor ontology (EFO) parent categories relevant to COVID-19 (respiratory disease, hematological or pulmonary function measurement,). Genes highlighted in bold indicate the loci involving COVID-19-relevant EFO categories with posterior probability for colocalization (PP4) > 0.5, suggesting evidence for shared genetic causality between eQTL and GWAS trait. In the *TLE* locus, the nearest genome-wide significant variant for forced expiratory volume in 1 second (FEV1) from Shrine et al.^68^ is more than 1Mb away, indicating that the association between the variant and FEV1 might be confounded by incomplete adjustment for height. **c-e**, Regional association plot for the GWAS signal on the upper panel and eQTL signal on the lower panel for *IFITM3* (c), *ERMP*1 (d), *MEPCE* (e) locus, where the eQTL for the corresponding gene colocalizes with the GWAS trait relevant to COVID-19. Genomic position of the variants is shown on the *x*-axis and -log10(*P*-value) of the GWAS or eQTL association on the *y*-axis. The lead GWAS and eQTL variants are highlighted.

In summary, our findings demonstrate replicable associations between current smoking, obesity, hypertension and increased bronchial epithelial *ACE2* expression potentially facilitating SARS-CoV-2 entry into host cells. While we do find evidence that the truncated ACE2 transcript is present in the bronchial epithelium, it does not appear to account for these associations. It is, however, likely that much of the inter-individual variation in COVID-19 is driven by a more complex molecular response to the virus in the airway epithelium than expression of *ACE2* alone. This supposition is supported by our results demonstrating that obesity, hypertension, cardiovascular comorbidities, as well as aging, are associated with a downregulation of mucosal immune response pathways similar to that seen in early SARS-CoV-2 infection in comparison to other viral infections. Together with clinical data and Mendelian randomization analyses of the causal role of smoking and BMI on severe COVID-19^47^, our result suggest that these important comorbidities increase COVID-19 susceptibility and severity by creating an airway microenvironment in which SARS-CoV-2 can gain a foothold before an effective host response is mounted. Conversely, we find that inflammatory airway conditions increase both innate and adaptive immune responses, potentially priming individuals for airway disease exacerbations in response to other viruses but not SARS-CoV-2, which has a different immune profile and does not appear to trigger exacerbations of airway diseases. Furthermore, we show that host genetics has a biologically meaningful effect on the expression of many genes that may play an important role in COVID-19, including genes of interest as future drug targets. We pinpoint multiple COVID-19-interacting genes for which genetic regulatory variants associate with immune- or respiratory-related outcomes; these variants may be candidates for host genetic risk factors for COVID-19, or its severity. Altogether, our findings of genetic and non-genetic factors affecting the expression of COVID-19-related genes in bronchial epithelium provide essential insights for understanding inter-individual variation of COVID-19 and developing therapeutic targets for COVID-19.

## Methods

### Study population

#### SubPopulations and InteRmediate Outcome Measures In COPD Study (SPIROMICS)

Data were obtained from participants who underwent research bronchoscopy within SPIROMICS, a multi-site prospective cohort study of participants ages 40-80, across four strata (never smokers, smokers without COPD, mild/moderate COPD, and severe COPD. Full SPIROMICS study details including inclusion and exclusion criteria have been previously published^13^. Participants enrolled in SPIROMICS who consented to a research bronchoscopy and met all local requirements (e.g. any laboratory tests that are required by institutional policy to be administered prior to a bronchoscopy) were deemed eligible. Additional exclusion criteria for the SPIROMICS bronchoscopy sub-study^48^ included history of cardiac disease or other comorbid condition severe enough to significantly increase risks based on investigator discretion, requirement of supplementary oxygen at rest based on arterial oxygen pressure less than 60 mm Hg or arterial oxygen saturation less than 88%, severe lung function impairment defined as post-bronchodilator forced expiratory volume in 1 second (FEV1) less than 30% predicted, and use of anticoagulation or antiplatelet therapies.

#### Severe Asthma Research Program (SARP)

Adult and pediatric patients with and without asthma were recruited to the SARP III cohort between Nov 1, 2012, and Oct 1, 2014, by seven clinical research centers in the USA. The SARP protocol is an ongoing, six-visit, 3-year, longitudinal cohort study in which 60% of participants have severe asthma as defined by the European Respiratory Society/American Thoracic Society (ERS/ATS) criteria^49^. A subset of participants underwent research bronchoscopy. Exclusion criteria included history of smoking (>5 pack year smoking history), co-existing lung disease, and uncontrolled comorbidities. All healthy control subjects had to have no history of asthma and normal lung function and methacholine bronchoprovocation testing. Participants with asthma had to meet ATS/ERS criteria for asthma (bronchodilator response to albuterol or positive methacholine bronchoprovocation test). Asthma had to be clinically stable at the time of bronchoscopy.

#### Mechanisms of ASThma Study (MAST)

Mild steroid naive asthmatics and healthy controls underwent research bronchoscopy. All healthy control subjects had to have no history of asthma or allergies. Participants with asthma had to have a positive methacholine bronchoprovocation test and could not have used steroids in 6 weeks prior to enrollment. Additional exclusion criteria included respiratory infection within 4 weeks of enrollment and pregnancy.

### Whole genome sequencing data

Trans-Omics for Precision Medicine (TOPMed) Project^14^ data freeze 9 consist of whole genome sequences of 160,974 samples with at least 15x average coverage, including 2,710 individuals from the SPIROMICS study. We obtained unphased genotypes for all individuals from the SPIROMICS study at sites with at least 10x sequencing depth (minDP10 call set) aligned to the human reference genome build GRCh38. Details regarding the DNA sample handling, quality control, library construction, clustering and sequencing, read processing, and sequence data quality control are described on the TOPMed website (*www.nhlbiwgs.org/genetic*). Variants passing all quality control (QC) filters were retained.

### Derivation of airway epithelial transcriptomic data in SPIROMICS, SARP, and MAST

Cytological brushings of the airway epithelium were obtained from lower lobe bronchi at the segmental or subsegmental carina. RNA was isolated with miRNeasy extraction kits (Qiagen Inc., Valencia, CA). RNA quantity and quality were evaluated using a NanoDrop Spectrophotometer (Thermo Fisher Scientific, Wilmington, DE) and Agilent 2100 Bioanalyzer (Agilent Technologies, Santa Clara, CA), respectively. Library preparation with multiplexing was performed using Illumina TruSeq Stranded Total RNA with Ribo-zero GOLD kit (SPIROMICS, SARP) or Human/Mouse/Rat kit (MAST) per manufacturer’s protocol. Samples were sequenced using one-hundred-fifty base-pair (SPIROMICS) or one-hundred base-pair (SARP, MAST) paired end reads via the Illumina HiSeq platform at the UCSF Sandler Genomics core. FASTQ files were quality filtered and aligned to the Ensembl GRCh38 genome build using STAR^50^. Read counts were normalized using the regularized logarithm transformation function of the DESeq2 package^51^ in R, and batch corrected using the Combat function in the SVA package^52^ in R. Outlying samples with low quality (low raw read counts, high percentage of reads mapped to multiple loci, high percentage of unmapped reads) were identified by hierarchical clustering and principal component analyses and excluded from the final datasets.

### Differential expression analysis of *ACE2* in relation to host/environmental factors

Visualization and analyses of single gene and gene signature analyses were done using RLE normalized and COMBAT batch corrected gene expression from the DESeq2 and SVA packages in R. Linear regression models were fitted to evaluate associations between *ACE2* expression (based on normalized count) and clinical variables. In SPIROMICS unadjusted models were evaluated along with models adjusted for potential confounders including smoking status, age, sex, body mass index (BMI), and race, as appropriate. For analyses of hypertension, adjusted analyses included anti-hypertensives as a covariate, in addition to sex, age, smoking status and race. In SARP adjusted analyses included covariates for asthma, steroid use (inhaled steroids alone or inhaled plus oral steroids), age, sex, race, and BMI, as appropriate. Analyses of hypertension were done only in participants with asthma (due to availability of data), but adjustments for sex, age, BMI, and race were still done. In MAST analyses were adjusted for age, sex, asthma disease status, race, and BMI.

### Differential exon usage

Following alignment, we indexed and sliced the SPIROMICS BAM files to include 51.6 kb of the *ACE2* genomic region (chrX:15,556,393-15,608,016 in the hg38 genome build) using samtools^53^. GTF files were manually curated to include the three exons that contribute to differential isoform expression of *ACE2*^24^. Full length *ACE2* transcripts are generated from two independent first exons, Exons 1a and 1b, with Exon 1b shared between these transcripts. The truncated *ACE2* transcript that does not bind the SARS-CoV-2 virus but is associated with an interferon-stimulated gene response in experimental models originates from Exon 1c. Coordinates from the pre-print manuscript by Onabajo, et al.^24^ were used to curate the GTF file: Exon 1a: chrX:15,601,956-15,602,158, Exon 1b: chrX:15,600,726-15,601,014, Exon 1c: chrX:15,580,281-15,580,420. The exons were then counted using the ASpli package^54^ in R. As per the ASpli and EdgeR package recommendations, raw exon counts were adjusted for gene counts to remove the signal from differential gene expression using the formula: (Exon Count in each sample*mean raw *ACE2* count)/raw *ACE2* gene count in that sample. To adjust for differences in sequencing depth between samples the transformed counts were then multiplied by the size factor variable generated by the DESeq2 package from the sequencing analysis. Linear models adjusting for batch were then used to analyze differences in exon usage in association with covariates of interest. The primary analysis was to evaluate whether *ACE2* exon 1c differential usage was associated with increases in our interferon-stimulated gene signature. In secondary analyses we determined whether clinical covariates were associated with differential exon 1c usage.

### Gene set enrichment analysis of expression changes induced by COVID-19

Differential expression and gene set enrichment analyses were done using the limma^55^ and FGSEA^28^ packages in R. Data underwent TMM normalization and the voom transformation followed by linear model fit with empirical Bayes moderation of the standard errors. We built COVID-19 relevant gene sets from publicly available differential gene expression data^27^ from participants who underwent nasal/oropharyngeal swab sampling at the time of acute respiratory illness for COVID-19 diagnosis (94 participants with COVID-19, 41 with other viral illness, 103 with no virus identified, viruses identified by metagenomic sequencing analysis). Expression gene sets were built using the 100 genes most up- and downregulated in association with infection type. Biological pathway gene sets were built by inputting the genes differentially downregulated between SARS-CoV-2 infection and other viral illness (*P* < 0.05) into the Ingenuity Pathway Analysis canonical pathway function. The pathway assessments were limited to downregulated genes given the relationship between downregulated gene sets and comorbidities in the initial analyses of the 100 gene sets. Gene set enrichment analyses were then performed using FGSEA^28^ and the CAMERA function^56^ in limma against gene lists ranked by their log fold change differential expression in association with comorbid clinical risk factors. Barcode plots were made using CAMERA. Normalized enrichment scores for heatmaps were extracted from FGSEA (not available through CAMERA, but CAMERA and FGSEA statistical results were similar). As smoking so strongly influences gene expression, in SPIROMICS differential expression analyses input into GSEA algorithms to evaluate clinical factors such as obesity, hypertension, cardiovascular conditions, and sex were first done in former smokers only to limit the effect of smoking, adjusting for age, sex and BMI if appropriate. In sensitivity analyses, we repeated the analyses in all subjects, adjusting for smoking status as well and found similar results. As asthma and steroid use so strongly influence gene expression, in SARP differential expression analyses of these other clinical factors were limited to asthma participants on inhaled but not oral steroids. Secondary analyses included all asthma participants adjusting for steroid use, with similar findings. Findings were considered significant at *P* < 0.05 and false discovery rate (FDR) < 0.05 if multiple corrections were necessary. For **Extended Data Fig. 7**, in which we evaluated COVID-19-related genes identified by experimental data from the SARS-CoV-2 *ex vivo* infection of primary human bronchial epithelial cells^31^ or thought to be proteins that interact with SARS-CoV-2^30^, we reported findings at the less stringent *P* < 0.05 as these analyses were hypothesis generating only.

### COVID-19-related genes

We mined the growing body of COVID-19 related literature to identify host genes implicated in SARS-CoV-2 infection discovered using different analytical approaches. The following studies were used to compose a list of COVID-19 candidate genes: 1) Hoffmann et al.^16^ that identified ACE2 as the receptor to be exploited by the SARS-CoV-2 for cellular entry, and proteases TMPRSS2 and cathepsin B/L both to be used by SARS-CoV-2 for S protein priming, whilst only TMPRSS2 is essential for viral entry and viral spread; 2) Gordon et al.^30^ that identified 332 high-confidence SARS-CoV-2-human protein-protein interactions; 3) Blanco-Melo et al.^31^ that explored the transcriptional response to SARS-CoV-2 *in vitro, ex vivo*, and *in vivo* models; 4) COVID-19 Cell Atlas (www.covid19cellatlas.org) that highlights 17 genes including cathepsins and other viral receptors or receptor associated enzymes; 5) Gassen et al.^32^ that showed the role of SARS-CoV-2 infection in restricting AMPK/mTORC1 activation and autophagy; 6) Wang et al.^29^ that reported a mediating role of CD147 (also known as BSG) in SARS-CoV-2 viral invasion.

To narrow the list of differentially expressed genes following SARS-CoV-2 infection from Blanco-Melo et al.^31^, we focused on the results from the *ex vivo* infection of primary human bronchial epithelial cells. To include in our candidate list, we chose genes that 1) have adjusted *P*-value < 0.05 in the differential expression analysis from primary cells and either cell lines (Calu-3 or ACE2-expressing A549 cells, low-MOI infection; excluded genes with adjusted *P* = 0) or samples derived from COVID-19 patients, and 2) log_2_ fold change > 0.5 in absolute scale in primary cells and log_2_ fold change > 1 in absolute scale in the other experiment.

In total, we selected 514 candidate genes implicated in COVID-19 from six different sources. Of them, 496 genes were expressed in bronchial epithelium.

### Expression quantitative trait mapping

Expression quantitative trait (eQTL) mapping was performed in 144 unrelated individuals from the SPIROMICS bronchoscopy sub-study with WGS genotype data from TOPMed and gene expression from bronchial epithelium profiled with RNA-seq following the analysis pipeline from the Genotype-Tissue Expression (GTEx) Consortium^15^. Gene expression data was normalized as follows: 1) read counts were normalized between samples using TMM^57^ with the edgeR package in R^58^, 2) genes with TPM ≥ 0.1 and unnormalized read count ≥ 6 in at least 20% of samples were retained, 3) expression values were transformed using rank-based inverse normal transformation across samples.

Next, Probabilistic Estimation of Expression Residuals^59^ (PEER) was used to infer hidden determinants of variability in gene expression levels due to technical and biological sources. According to the optimization analysis for selection of PEERs by sample size to maximize *cis*-eGene discovery done in GTEx^15^, 15 PEERs were chosen to be used as covariates in eQTL mapping together with 4 genotype PCs and sex imputed from genotype data.

To control population structure, principal component analysis (PCA) was conducted on post-variant QC genotype data from unrelated SPIROMICS individuals. More precisely, PCA was performed on a set of LD-independent autosomal biallelic single nucleotide polymorphisms from not long-range LD regions^60^ with a call rate ≥ 99% and MAF ≥ 0.05 using smartpca from

EIGENSTRAT (*github.com/argriffing/eigensoft/tree/master/EIGENSTRAT*), turning off outlier removal (option `-m 0`). LD pruning was performed using Plink 1.9^61^ based on pairwise genotypic correlation of 200 SNPs at a time, with a step of 100 SNPs, and using LD threshold of > 0.1 to remove one of a pair of SNPs (option `--indep-pairwise 200 100 0.1`). Top 4 PCs were chosen to be used to correct for population stratification. The top 4 PCs explained > 0.1% of the variance, and were associated with subpopulations inferred from 1000 Genomes Project using k-nearest neighbors clustering (F-test *P* < 2×10^−10^, adj R^2^ = 0.36 − 0.98).

*Cis*-eQTL mapping was performed using tensorQTL^62^ across 22,738 genes and 6,605,907 variants with MAF ≥ 0.05 and variant call rate ≥ 0.9. Window-size was set to 1 Mb from the transcription start site (TSS) of the gene according to the GENCODE version 33, 10,000 permutations were used to correct for multiple testing, and false discovery rate (FDR) < 0.05 was used to identify genes with statistically significant eQTLs (eGenes). We also used tensorQTL to map conditionally independent *cis*-eQTLs.

Lead *cis*-eQTL effect size was quantified as allelic fold change^63^ (aFC), ratio of expression of the haplotype carrying the alternative allele to expression of the haplotype carrying the reference allele of an eQTL. Gene expression data normalized with DESeq2 size factors^51^ and log_2_-transformed were used as input together with the processed genotype VCF file. aFC was calculated requiring at least 2 samples (`--min_samps 2`) and minimum 1 observation of each allele (`--min_alleles 1`), and adjusting for the same covariates as in *cis*-eQTL mapping. To calculate confidence intervals 100 bootstrap samples were used. aFC estimates that hit the absolute cap value (log_2_(100)) were set to missing.

### Cell type interacting expression quantitative trait mapping

Firstly, we used xCell^64^ to estimate 64 immune and stroma cell types from the gene expression signatures of bronchial epithelium. TPM expression matrix of 144 bronchial brush samples together with 30 samples from each tissue type from GTEx was uploaded to the UCSF xCell Webtool (*xcell.ucsf.edu*). Then, the following linear regression model was used to map cell type interaction eQTLs (ieQTLs): p ∼ g+i+g×i+C, where *p* is the phenotype vector (inverse normalized gene expression), *g* is the genotype vector, *i* is the cell type enrichment score from xCell (inverse normalized), *g×i* is the interaction term, and *C* is the covariates matrix as used in eQTL mapping. Cell types with a median xCell enrichment score > 0.05 were included in the analysis. There were 29 cell type signatures that met this criteria: B cells, basophils, CD4 TCM, CD4 TEM, CD8 naïve T cells, common lymphoid progenitor (CLP), common myeloid progenitor (CMP), class switched memory B cells, DCs, eosinophils, HSCs, keratinocytes, M1 and M2 macrophages, monocytes, osteoblasts, plasma cells, preadipocytes, sebocytes, smooth muscle, TH2 cells, antigen-presenting, immature, classical and plasmacytoid dendritic cells, pro B cells, and microenvironment and immune scores. Mapping of ieQTLs was done using tensorQTL^62^, and only variants within 1Mb of the TSS of each gene tested and with MAF > 0.1 in the samples belonging to the top and bottom halves of the distribution of cell type abundance were included. Regression coefficients and *P*-values were calculated for all terms in the model, and ieQTLs were identified by testing for the significance of the interaction term. Top nominal *P*-values for each gene were corrected for multiple testing at the gene level using eigenMT^65^ as implemented in tensorQTL. Significance across genes was determined by adjusting eigenMT *P*-values using the Benjamini-Hochberg procedure with FDR of 0.05.

### Replication of *cis*-eQTLs in GTEx

We performed replication of *cis*-eQTLs (gene-variant pairs) found from bronchial epithelium in the Genotype-Tissue Expression (GTEx) project v8 release^15^. Using *cis*-eQTL summary statistics across 49 tissues from GTEx, we calculated the proportion of true positives^66^, π_1_, to estimate the proportion of sharing of *cis*-eQTLs between bronchial epithelium and GTEx tissues. We assessed the allelic direction of the *cis*-eQTLs from bronchial epithelium and GTEx tissues by calculating concordance rate, the proportion of gene-variant pairs with the same allelic direction. This comparison was restricted to *cis*-eQTLs with nominal *P*-value < 1⨯10^-4^ in the given GTEx tissue.

Next, we analyzed the replication and concordance measure as a function of sample size and median cell type enrichment scores for seven cell types^36^: adipocytes, epithelial cells, hepatocytes, keratinocytes, myocytes, neurons, and neutrophils. Tissues with median enrichment score > 0.1 were classified as being enriched for the given cell type. We used Wilcoxon rank sum test to estimate the difference in replication and concordance estimates between tissues enriched or not enriched for the given cell type.

### *cis*-eQTLs not identified in previous large eQTL catalogs

To investigate the tissue-specificity of *cis*-eQTLs from bronchial epithelium, we performed gene-level lookup in GTEx v8 and eQTLGen Consortium^35^. We identified genes with significant regulatory effects in SPIROMICS (FDR < 0.05) that were tested in neither catalog.

Then, we used the functional profiling webtool g:GOSt (version e99_eg46_p14_f929183) from g:Profiler^67^ to perform pathway analysis of the 492 significant eGenes in SPIROMICS not tested in GTEx v8 Lung. Method g:SCS was used for multiple testing correction corresponding to experiment-wide threshold of *α* = 0.05. Significant eGenes from SPIROMICS (n = 4,881) that have at least one annotation (option “Custom over annotated genes”) were used as a background in the enrichment test.

### pheWAS of lead COVID-19 *cis*-eQTLs in SPIROMICS

We performed phenome-wide association studies in 1,980 non-Hispanic White (NHW) and 696 individuals from other ethnic and racial groups from SPIROMICS for the 108 lead *cis*-eQTLs to evaluate for phenotypic associations with spirometric measures, cell count differentials, immunoglobulin concentrations, longitudinal exacerbation risk, self-reported asthma history, cardiovascular diseases, CT scan measures of emphysema (bilateral percentage lung density <-950HFU at total lung capacity), CT scan functional small airways disease (PRM-fSAD), and alpha1-antitrypsin concentrations (subgroup of 1,191 NHW and 396 from other racial/ethnic groups). PheWAS regression-based models were performed using PLINK 2/0 and included principle components of ancestry, sex, BMI, age, and smoking pack-years. Models for CT scan measures also included site and height while alpha1-antitrypsin concentrations included c-reactive protein.

CT scan measures, eosinophil counts, and IgE concentrations were log-transformed. Significance threshold was set for the number of eQTLs tested across phenotypes (*P* < 4.63⨯10^-4^).

### Lookup of phenome-wide associations with PhenoScanner v2

PhenoScanner v2^37,38^ was used to lookup phenotype associations for the *cis*-eQTL variants from large-scale genome-wide association studies (GWAS) with association *P*-value < 10^-5^. We queried PhenoScanner database based on the rs IDs of the lead *cis*-eQTLs obtained from dbSNP version 151 (GRCh38p7, including also former rs ID to query). The phenoscanner R package (*github.com/phenoscanner/phenoscanner*) was used to perform the queries. Query results were filtered to keep one association for each of the variants per trait, preferring summary statistics from newer studies, studies with larger sample size, or based on UK Biobank data (GWAS round 1 results from the Neale Lab). Description of Experimental Factor Ontology (EFO) terms and classification to EFO broader categories were obtained from the GWAS Catalog or by manually searching EMBL-EBI EFO webpage (*www.ebi.ac.uk/efo/*).

The regulatory variants for *CEP250, FAR2*, and *TLE3* have phenotypic associations with both body height and pulmonary function test (PFT) measures from PhenoScanner. As GWAS analyses from the Neale Lab using UK Biobank data do not include height as a covariate in the model, we used the results of the lung function GWAS by Shrine et al.^68^ to confirm if the suggestive signal for PFT trait has been observed before or rather seems to be an artefact of incomplete adjustment for height. Of note, Shrine et al.^68^ have discovered 279 lung functions signals in the meta-analysis of UK Biobank and the SpiroMeta Consortium. We looked up the nearest GWAS hits to the eQTL, and calculated LD between the variants in the African and European populations using LDpop^69^ web tool.

### Colocalization analysis

Multiple trait associations observed for a single variant do not necessarily translate into shared genetic causality. To assess evidence for shared causal variant of a *cis*-eQTL and a GWAS trait, we used the Bayesian statistical test for colocalization, coloc. We used the newer version of coloc^70^ that allows conditioning and masking to overcome one single causal variant assumption (condmask branch of coloc from *github.com/chr1swallace/coloc*). We only tested colocalization for loci where the eQTL had at least one phenotypic association based on the lookup analysis with Phenoscanner from the following EFO parent categories: hematological measurement, pulmonary function measurement, respiratory disease. From each of the smaller EFO categories, we chose one trait with the smallest *P*-value for which we were able to find summary statistics using GWAS Catalog REST API or among the Neale Lab GWAS round 2 results (*www.nealelab.is/uk-biobank/*). Coloc was run on a 500-kb region centered on the lead *cis*-eQTL (+/- 250 kb from the variant) with priors set to *p*_*1*_ = 10^-4^, *p*_*2*_ = 10^-4^, *p*_*3*_ = 5×10^-6^. We used the coloc.signals() function with `mode = iterative` and `method = mask` for GWAS traits with LD data from the 1000 Genomes Project to match the ancestry of the discovery population (*e.g*., choosing CEU for LD if the discovery population is of European ancestry). We allowed for a maximum of three variants to mask, with an r^2^ threshold of 0.01 to call two signals independent and *P*-value threshold of 1⨯10^-5^ to call the secondary signal significant. We used `method = single` for the eQTLs, because the corresponding eGenes did not have secondary independent signals. We prioritized eGenes with posterior probability for colocalization (PP4) > 0.5 as loci with evidence for colocalization.

## Data availability

The SPIROMICS gene expression data will be available in dbGaP. The SARP gene expression data will be available through the Gene Expression Omnibus (GEO). The MAST dataset is available in GEO under accession number GSE67472. TOPMed WGS freeze 9 data are available in dbGaP under accession number phs001927.

Full eQTL summary statistics for the 496 COVID-19-related genes can be downloaded from https://github.com/LappalainenLab/spiromics-covid19-eqtl/tree/master/eqtl/summary_stats.

## Code availability

eQTL mapping analyses code has been deposited to the GitHub repository at https://github.com/LappalainenLab/spiromics-covid19-eqtl.

## Data Availability

The SPIROMICS gene expression data will be available in dbGaP. The SARP gene expression data will be available through the Gene Expression Omnibus (GEO). The MAST dataset is available in GEO under accession number GSE67472. TOPMed WGS freeze 9 data are available in dbGaP under accession number phs001927.
Full eQTL summary statistics for the 496 COVID-19-related genes can be downloaded from https://github.com/LappalainenLab/spiromics-covid19-eqtl/tree/master/eqtl/summary_stats.

## Acknowledgements

This work was funded by following funding sources: R01HL142992 (V.E.O.), R01HL137880 (V.E.O.), F30HG011194 (M.M.), R01MH106842 (T.L.), R01HL142028 (T.L., R.G.B. and S.K.), R01GM122924 (T.L.), UM1HG008901 (T.L.), R01GM124486 (T.L.), K23HL123778 (S.A.C.), R01HL121774 (S.A.C.), U01HL137880 (S.A.C.).

Molecular data for the Trans-Omics in Precision Medicine (TOPMed) program was supported by the National Heart, Lung and Blood Institute (NHLBI). Genome Sequencing for “NHLBI TOPMed: SubPopulations and InteRmediate Outcome Measures In COPD Study” (phs001927) was performed at the Broad Institute Genomics Platform (HHSN268201600034I). Core support including centralized genomic read mapping and genotype calling, along with variant quality metrics and filtering were provided by the TOPMed Informatics Research Center (3R01HL-117626-02S1; contract HHSN268201800002I). Core support including phenotype harmonization, data management, sample-identity QC, and general program coordination were provided by the TOPMed Data Coordinating Center (R01HL-120393; U01HL-120393; contract HHSN268201800001I). We gratefully acknowledge the studies and participants who provided biological samples and data for TOPMed. A list of banner authors for the NHLBI Trans-Omics for Precision Medicine (TOPMed) Consortium is provided in the **Supplementary Note**.

The authors thank the SPIROMICS participants and participating physicians, investigators and staff for making this research possible. More information about the study and how to access SPIROMICS data is available at www.spiromics.org. The authors would like to acknowledge the University of North Carolina at Chapel Hill BioSpecimen Processing Facility for sample processing, storage, and sample disbursements (http://bsp.web.unc.edu/). We would like to acknowledge the following current and former investigators of the SPIROMICS sites and reading centers: Neil E Alexis, MD; Wayne H Anderson, PhD; Mehrdad Arjomandi, MD; Igor Barjaktarevic, MD, PhD; R Graham Barr, MD, DrPH; Patricia Basta, PhD; Lori A Bateman, MSc; Surya P Bhatt, MD; Eugene R Bleecker, MD; Richard C Boucher, MD; Russell P Bowler, MD, PhD; Stephanie A Christenson, MD; Alejandro P Comellas, MD; Christopher B Cooper, MD, PhD; David J Couper, PhD; Gerard J Criner, MD; Ronald G Crystal, MD; Jeffrey L Curtis, MD; Claire M Doerschuk, MD; Mark T Dransfield, MD; Brad Drummond, MD; Christine M Freeman, PhD; Craig Galban, PhD; MeiLan K Han, MD, MS; Nadia N Hansel, MD, MPH; Annette T Hastie, PhD; Eric A Hoffman, PhD; Yvonne Huang, MD; Robert J Kaner, MD; Richard E Kanner, MD; Eric C Kleerup, MD; Jerry A Krishnan, MD, PhD; Lisa M LaVange, PhD; Stephen C Lazarus, MD; Fernando J Martinez, MD, MS; Deborah A Meyers, PhD; Wendy C Moore, MD; John D Newell Jr, MD; Robert Paine, III, MD; Laura Paulin, MD, MHS; Stephen P Peters, MD, PhD; Cheryl Pirozzi, MD; Nirupama Putcha, MD, MHS; Elizabeth C Oelsner, MD, MPH; Wanda K O’Neal, PhD; Victor E Ortega, MD, PhD; Sanjeev Raman, MBBS, MD; Stephen I. Rennard, MD; Donald P Tashkin, MD; J Michael Wells, MD; Robert A Wise, MD; and Prescott G Woodruff, MD, MPH. The project officers from the Lung Division of the National Heart, Lung, and Blood Institute were Lisa Postow, PhD, and Lisa Viviano, BSN; SPIROMICS was supported by contracts from the NIH/NHLBI (HHSN268200900013C, HHSN268200900014C, HHSN268200900015C, HHSN268200900016C, HHSN268200900017C, HHSN268200900018C, HHSN268200900019C, HHSN268200900020C), grants from the NIH/NHLBI (U01 HL137880 and U24 HL141762), and supplemented by contributions made through the Foundation for the NIH and the COPD Foundation from AstraZeneca/MedImmune; Bayer; Bellerophon Therapeutics; Boehringer-Ingelheim Pharmaceuticals, Inc.; Chiesi Farmaceutici S.p.A.; Forest Research Institute, Inc.; GlaxoSmithKline; Grifols Therapeutics, Inc.; Ikaria, Inc.; Novartis Pharmaceuticals Corporation; Nycomed GmbH; ProterixBio; Regeneron Pharmaceuticals, Inc.; Sanofi; Sunovion; Takeda Pharmaceutical Company; and Theravance Biopharma and Mylan.

## Author contributions

S.K., T.L., S.A.C., P.G.W. and V.E.O. designed the study. S.A.C., S.G., J.N. and S.N. performed differential expression analyses. S.K., M.M., A.P. and K.L.B. performed eQTL analyses. V.E.O. and E.A. performed phenome-wide association analysis. I.Z.B., R.G.B., E.R.B., R.P.B., A.P.C., C.B.C., D.J.C., G.J.C., J.L.C., M.K.H., N.N.H., E.A.H., R.J.K., J.A.K., F.J.M., M.N.M., D.A.M., R.P., S.P.P., M.C., L.C.D., S.C.E., J.V.F., E.I., N.N.J., B.D.L., X.L., W.C.M., S.E.W., J.Z. and C.L. were involved in data collection and processing. T.L. and S.A.C. supervised the work. S.K., T.L. and S.A.C. wrote the first draft. J.L.C., R.P.B., R.P., S.E.W., R.J.K., A.P.C., M.M. and K.L.B. contributed to editing of the manuscript. All authors approved the final version of the manuscript.

