## Extended Data Figures for "Genetic and non-genetic factors affecting the expression of COVID-19 relevant genes in the large airway epithelium"

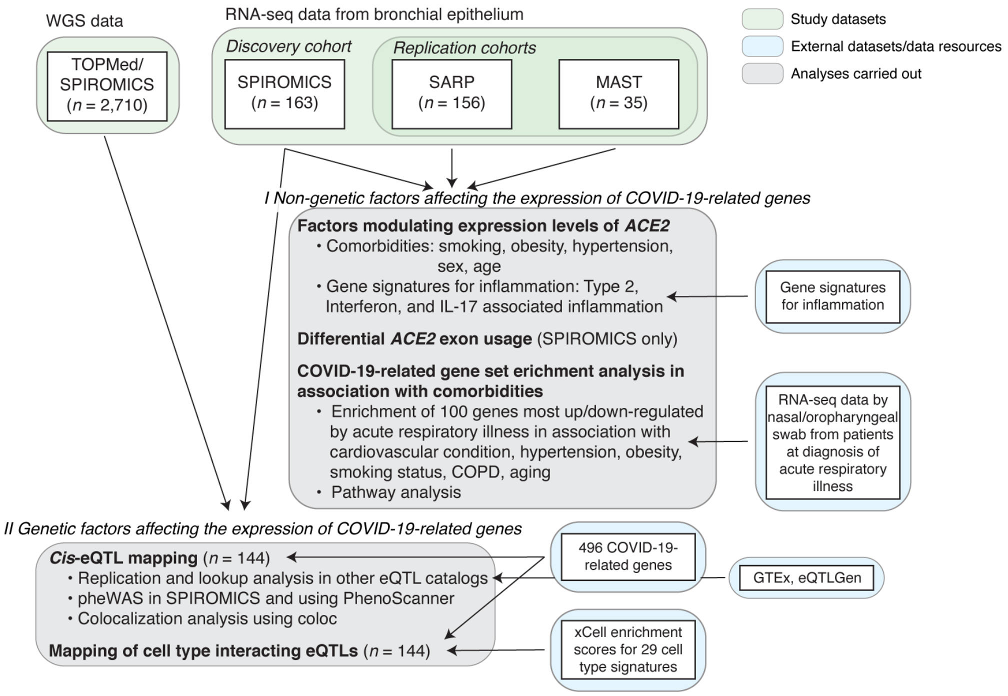


**Extended Data Fig. 1. Study design.** Graphical illustration of analyses (grey boxes) carried out to study non-genetic and genetic factors affecting the expression of COVID-19-related genes in bronchial epithelium. Input datasets for these analyses are denoted with a green box (WGS and RNA-seq) and external datasets or data resources used in these analyses are denoted with a blue box.

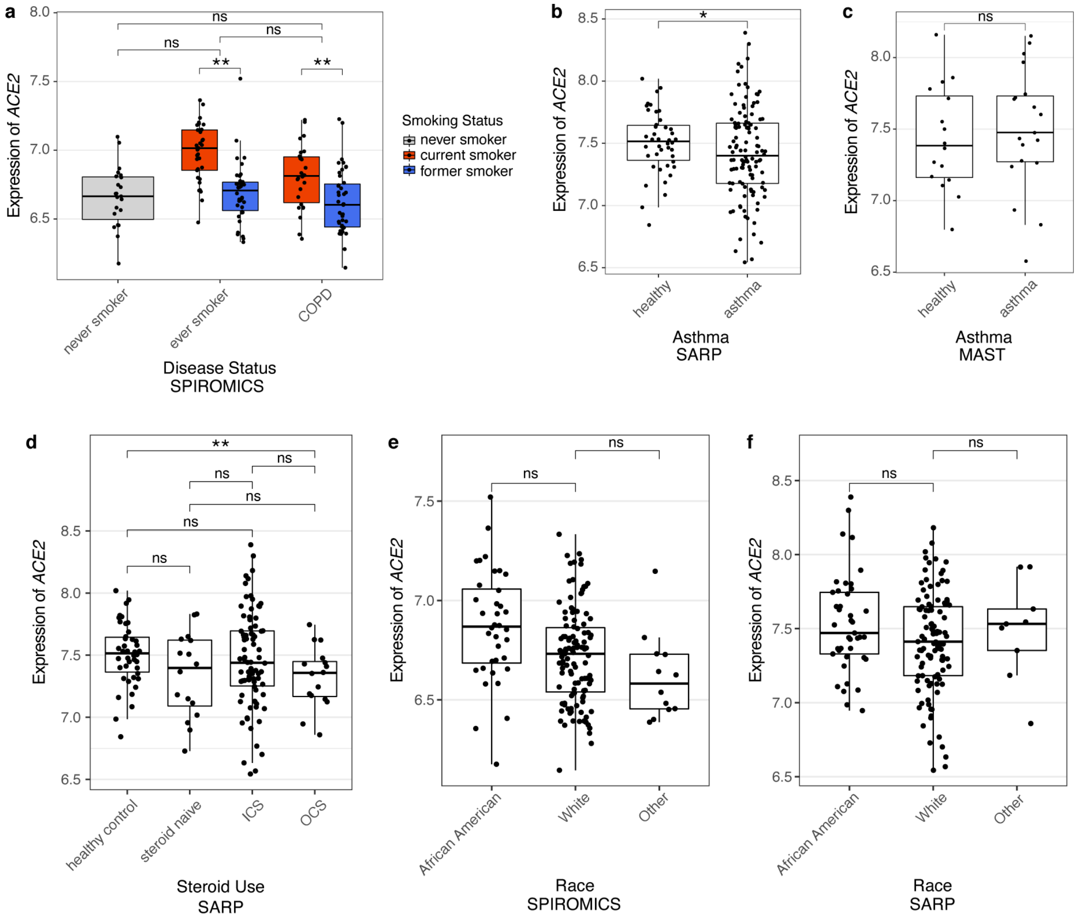


**Extended Data Fig. 2. Associations between *ACE2* gene expression and COPD, asthma, steroid use, and race. a**, Boxplots showing *ACE2* $log_2$ gene expression in association with COPD in SPIROMICS. While current smoking significantly affected *ACE2* $log_2$ gene expression, there was no association with COPD after adjusting for smoking status (*P* > 0.05 for never smokers compared to current/former smokers with COPD and P > 0.05 for current/former smokers without COPD vs current/former smokers with COPD). **b-c**, Boxplots showing *ACE2* $log_2$ gene expression in association with asthma in SARP and MAST. In SARP, *ACE2* levels were slightly lower in asthma compared to healthy controls. There was no significant association in MAST. **d**, When stratified by any steroid use in *SARP*, *ACE2* levels were significantly lower in asthmatics on oral but not inhaled steroids compared to healthy controls. ICS - inhaled steroids, OCS - oral steroids. **e-f**, Boxplots showing *ACE2* $log_2$ gene expression in association with race in SPIROMICS and SARP. While African American race was associated with increased *ACE2* expression in both SPIROMICS (*P* = 0.002 before adjustments) and SARP (*P* = 0.042 before adjustment), this association was non-significant after adjusting for covariates. *P*-values: ****<0.0001, ***<0.001, **<0.01, *<0.05, ns=not significant in linear models adjusted for covariates.

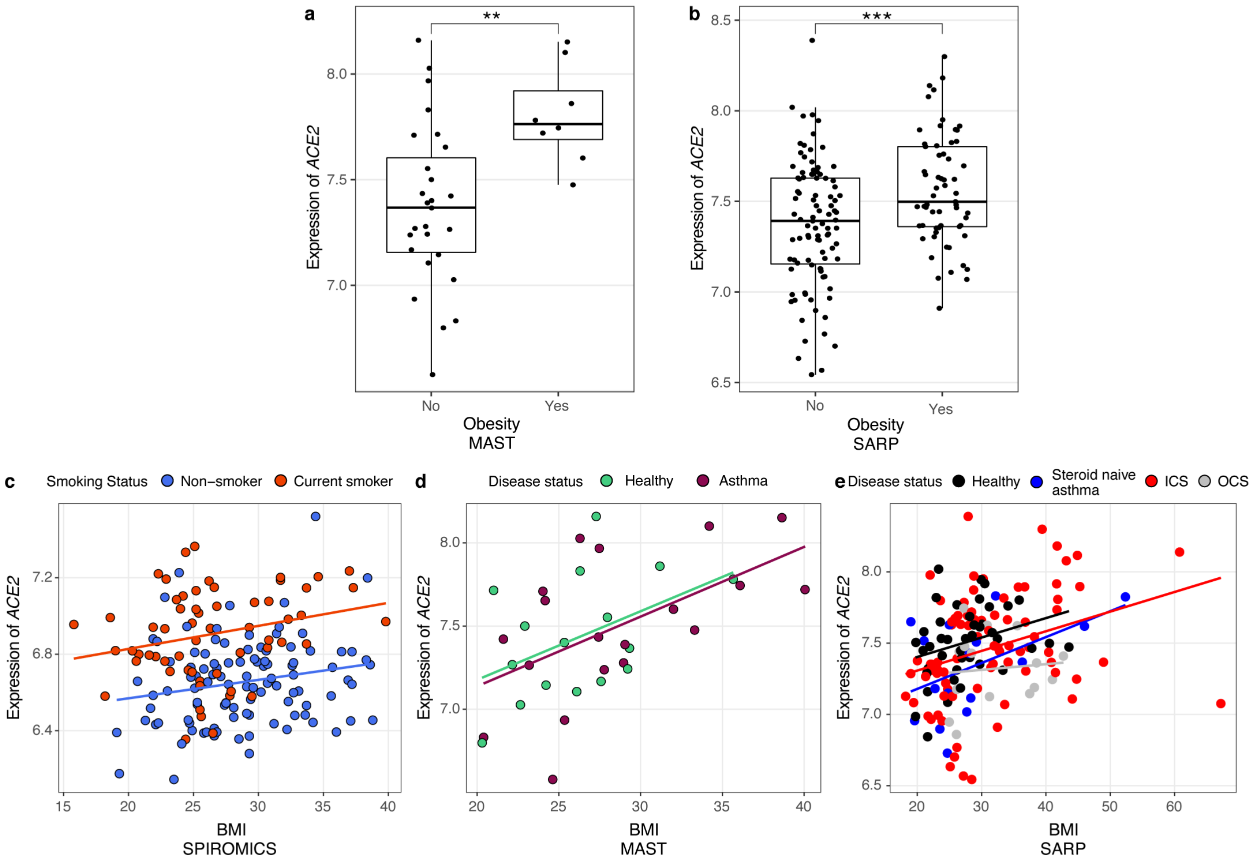


**Extended Data Fig. 3. Associations between *ACE2* gene expression and obesity. a-b**, Boxplots showing *ACE2* $\log_2$ gene expression increases with obesity across MAST and SARP. *P*-values: ****<0.0001, ***<0.001, **<0.01, *<0.05, ns=not significant in linear models adjusted for covariates. **c-e**, Scatterplots showing *ACE2* $\log_2$ gene expression also increase with increasing body mass index as a continuous variable. Plots show that *ACE2* expression increases across both non- and current smokers in SPIROMICS (**c**), healthy controls and asthma in MAST (**d**), and healthy controls, steroid naive asthma, and asthma on inhaled steroids in SARP (**c**).

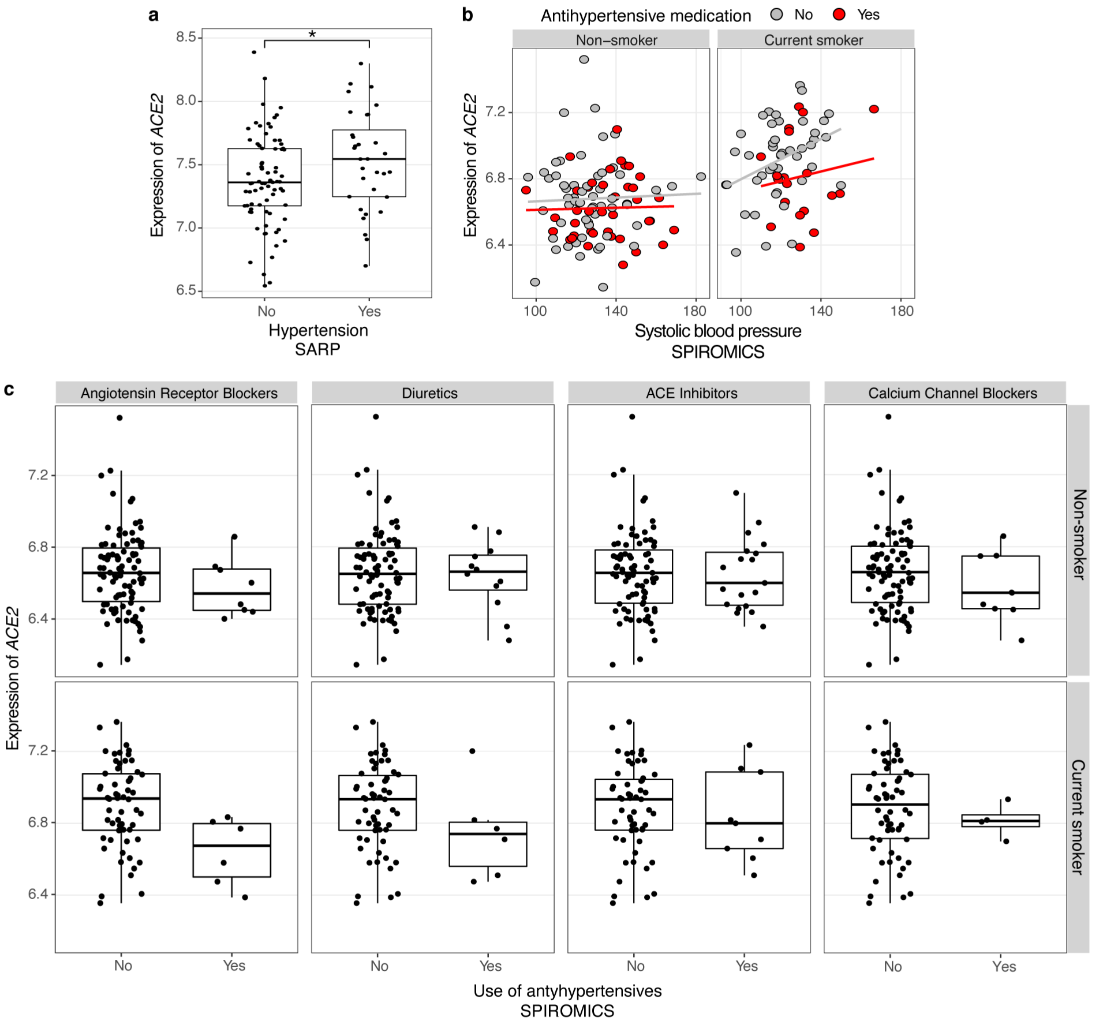


**Extended Data Fig. 4. Associations between *ACE2* gene expression and hypertension, and use of antihypertensives. a**, Boxplots showing *ACE2* $\log_2$ gene expression increases with hypertension in SARP, data not collected in MAST. *P*-values: ****<0.0001, ***<0.001, **<0.01, *<0.05, ns=not significant in linear models adjusted for covariates. **b**, Scatterplots showing *ACE2* $\log_2$ gene expression also increase when considering systolic blood pressure as a continuous variable among current smokers. **c**, Boxplots showing *ACE2* $\log_2$ gene expression in association with the use of antihypertensives by antihypertensive class (Angiotensin Receptor Blockers, Diuretics, ACE Inhibitors, Calcium Channel Blockers, respectively). Use of Angiotensin Receptor Blockers (ARBs) and Diuretics are associated with lower ACE2 levels in unadjusted model (*P* = 0.021) and adjusted model in current smokers only (*P* = 0.023).

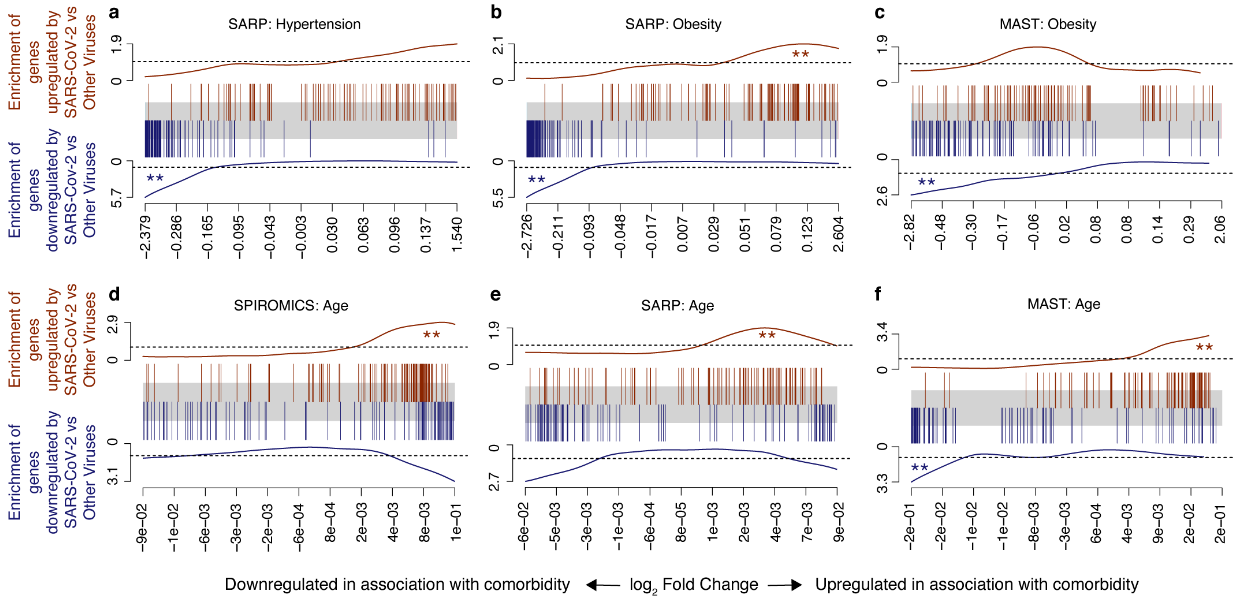


**Extended Data Fig. 5. COVID-19 and other viral illness related gene set enrichment analyses in association with comorbidities in SPIROMICS, SARP, and MAST. a-f**, Barcode plots in which the vertical lines represent the 100 genes most upregulated (red) or downregulated (blue) in nasal/oropharyngeal swab samples obtained from patients with COVID-19 as compared other viruses at the time of diagnosis of an acute upper respiratory infection. These gene sets are plotted against log fold gene expression changes arranged from most downregulated to most upregulated with a given comorbidity (horizontal grey bar). Lines above (red) and below (blue) the bar represent the running sum statistic with a significant finding indicated when the line crosses the dashed line at either end of the plot. Comorbidities included in the analyses are hypertension in SARP (**a**), obesity in SARP and MAST (**b-c**), age in SPIROMICS, SARP, and MAST (**d-f**).

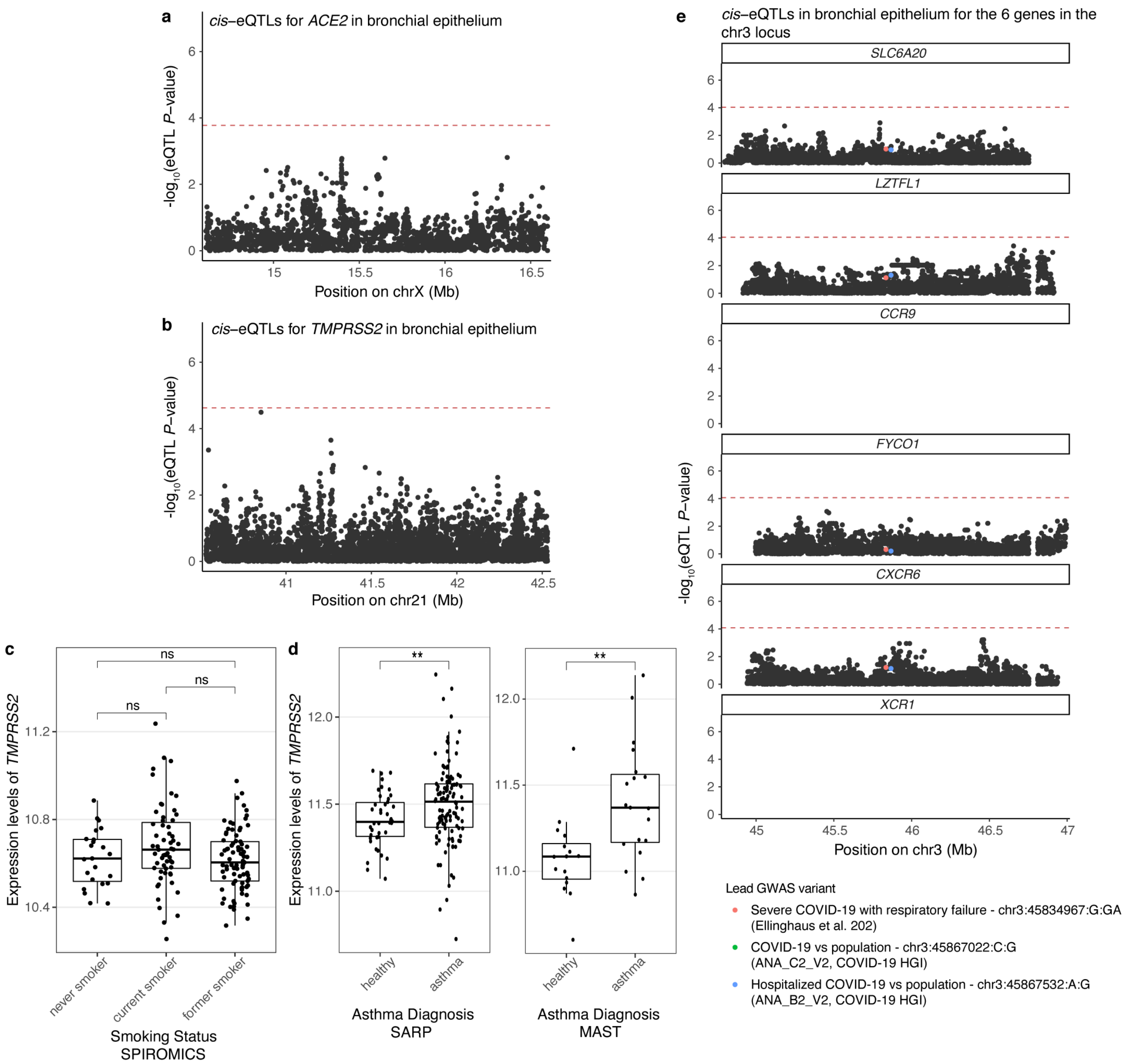


**Extended Data Fig. 6. Regulatory genetic effects of *ACE2, TMPRSS2,* and six genes in the chr3 cluster associated with COVID-19 in bronchial epithelium, and the effect of smoking of *TMPRSS2*.** **a-b,** Regional association plot illustrating no statistically significant regulatory effects on the gene expression levels of *ACE2* (**a**) and *TMPRSS2* (**b**). Each data point represents a genetic variant in the +/-1Mb *cis*-window of the transcription start site of the given gene. Red dashed line denotes the gene-level significance threshold for the given gene. **c**, Boxplot showing *TMPRSS2* $\log_2$ gene expression in association with smoking in SPIROMICS. Smoking has a more modest effect on the expression of *TMRPSS2* and is only significantly increased in current smokers when compared to never and current smokers overall ($P = 0.026$) but not when stratified by former and never smoker, as shown in the boxplot. ns=not significant in linear models adjusted for covariates. **d**, Boxplot showing *TMPRSS2* $\log_2$ gene expression in association with asthma in SARP (left) and MAST (right). Expression levels of *TMPRSS2* are higher in asthmatic than healthy controls ($P = 0.038$ in SARP, $P = 0.0028$ in MAST). **e,** Regional association plots as shown on **a** illustrating no statistically significant regulatory effects on the gene expression levels of *SLC6A20, LZTFL1, FYCO1, CXCR6* in the chr3 locus associated with hospitalized COVID-19 (COVID-19 HGI[9]) and severe COVID-19 with respiratory failure (Ellinghaus et al.[8]). *CCR9* and *XCR1*, were not expressed in our bronchial epithelium dataset.

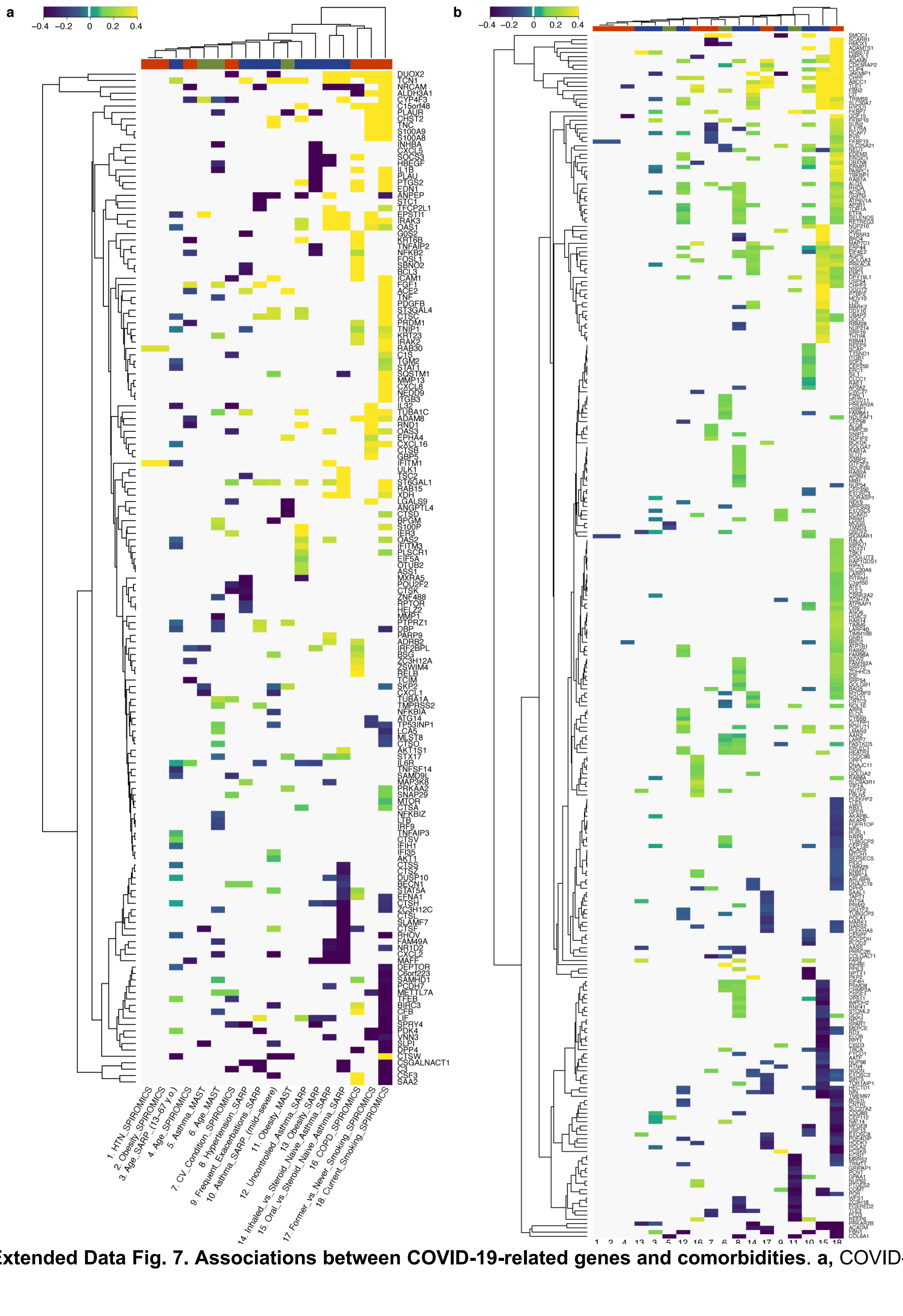


**Extended Data Fig. 7. Associations between COVID-19-related genes and comorbidities. a,** COVID-

19-related genes identified in experimental models with primary human bronchial epithelial cells from Blanco-Melo et al.[31] are shown in the heatmap (rows) plotted against comorbidities (columns) with genes differentially down- and upregulated in association with that comorbidity at *P*-value < 0.05 indicated in blue and yellow, respectively. **b,** Similar heatmap showing genes identified as potentially interacting with the SARS-CoV-2 virus by protein-protein interaction analyses (rows) from Gordon et al.[30]. All genes not associated at *P*-value < 0.05 were shrunk to zero (white). Euclidean distance with average linkage was used for clustering.

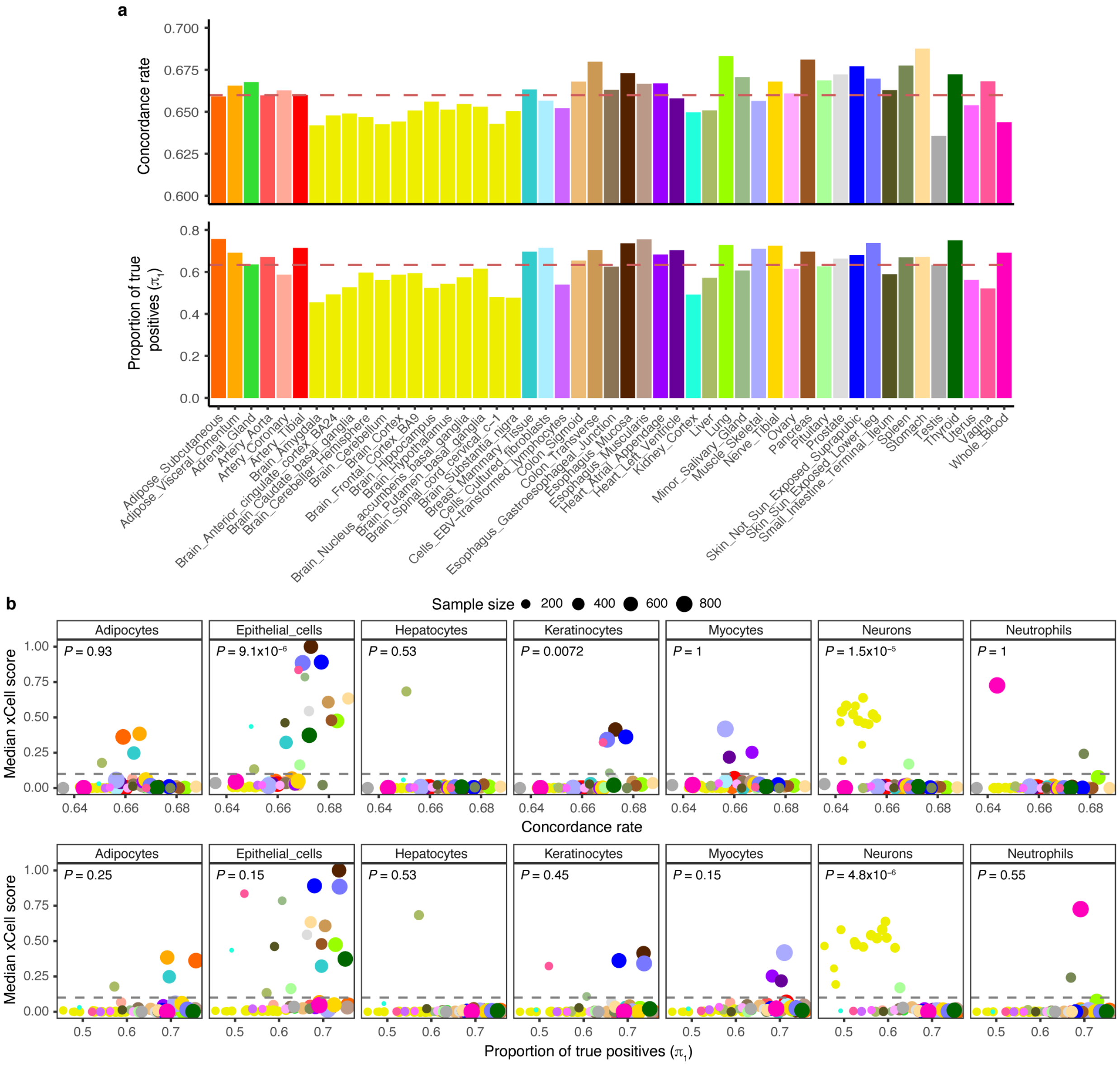


**Extended Data Fig. 8. Replication of *cis*-eQTLs in GTEx. a**, Replication of *cis*-eQTLs from bronchial epithelium in GTEx v8 using the concordance rate (proportion of gene-variant pairs with the same direction of the effect, upper panel) and proportion of true positives ($\boldsymbol{\pi}_1$, lower panel). Red dashed line denotes the median of the respective replication measure. **b**, Relationship between the two replication measures as the function of cell type enrichment of the tissues measured as median enrichment score from xCell for seven cell types: adipocytes, epithelial cells, hepatocytes, keratinocytes, myocytes, neurons, and neutrophils. Grey dashed line denotes median enrichment score > 0.1, which classifies tissues as enriched for the given cell type or not. Wilcoxon rank sum test was used to estimate the difference in replication estimates between tissues enriched or not enriched for the given cell type.

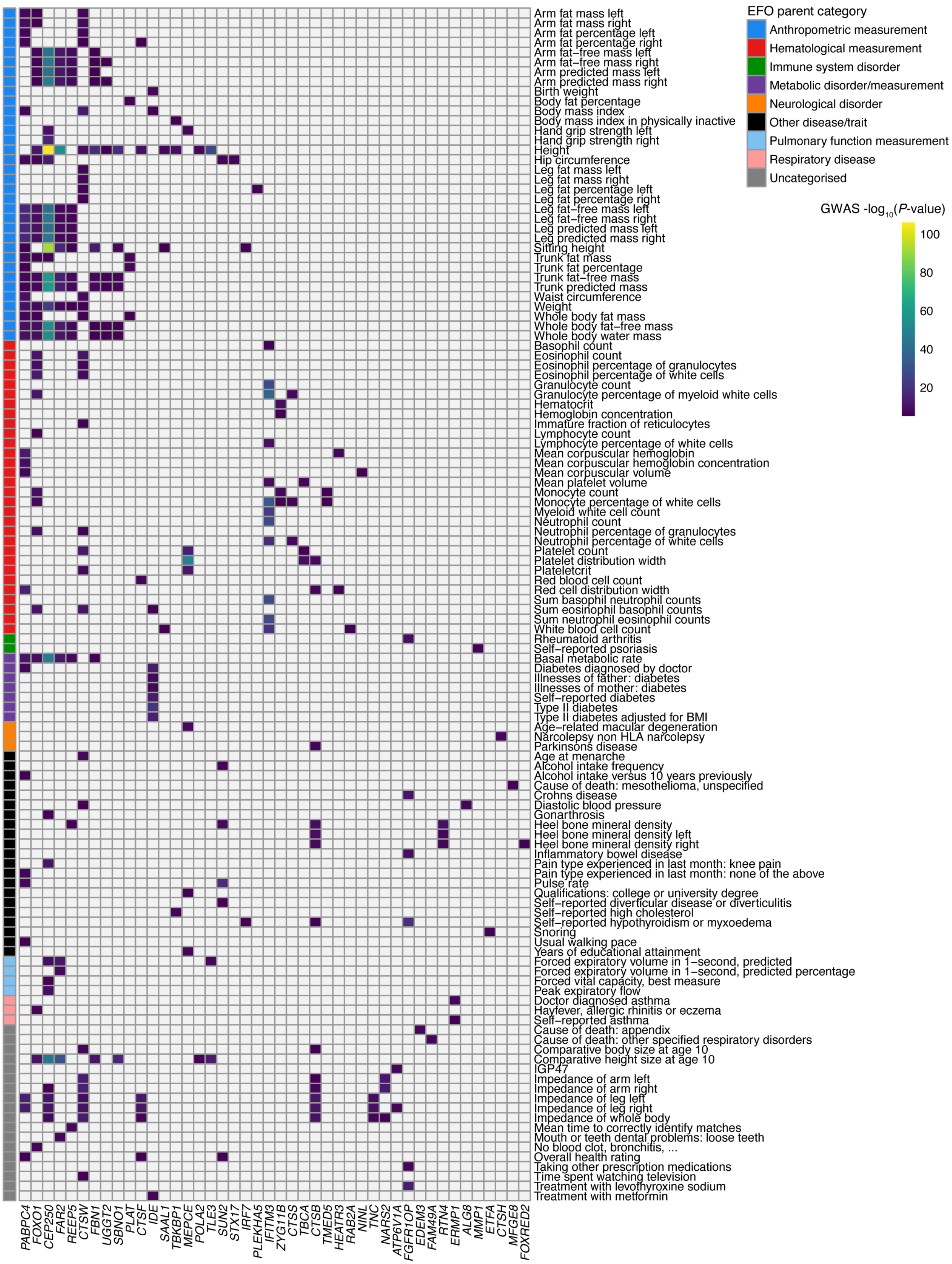


**Extended Data Fig. 9. PheWAS associations for the 44 out of 108 lead *cis*-eQTLs associated with**

**COVID-19-related genes by Phenoscanner v2.** Heatmap showing the pheWAS associations with the eQTLs for COVID-19-related genes. Traits are grouped based on the experimental factor ontology (EFO) terms, and EFO terms with few traits grouped into one group, “Other diseases/traits”. Tiles are colored by the $-\log_{10}$(GWAS *P*-value) or grey, if there was no suggestive signal obtained for the given variant and trait (GWAS *P*-value > $10^{-5}$).
