## Supplementary Note for "Genetic and non-genetic factors affecting the expression of COVID-19 relevant genes in the large airway epithelium"

**Supplementary Note.** NHLBI Trans-Omics for Precision Medicine (TOPMed) Consortium Banner Authorship List.

Namiko Abe[1], Gonçalo Abecasis[2], Francois Aguet[3], Christine Albert[4,5], Laura Almasy[6], Alvaro Alonso[7], Seth Ament[8], Peter Anderson[9], Pramod Anugu[10], Deborah Applebaum-Bowden[11], Kristin Ardlie[3], Dan Arking[12], Donna K Arnett[13], Allison Ashley-Koch[14], Stella Aslibekyan[15], Tim Assimes[16], Paul Auer[17], Dimitrios Avramopoulos[12], John Barnard[18], Kathleen Barnes[19], R. Graham Barr[20], Emily Barron-Casella[12], Lucas Barwick[21], Terri Beaty[12], Gerald Beck[22], Diane Becker[23], Lewis Becker[12], Rebecca Beer[24], Amber Beitelshees[8], Emelia Benjamin[25,26], Takis Benos[27], Marcos Bezerra[28], Larry Bielak[2], Joshua Bis[29], Thomas Blackwell[2], John Blangero[30], Eric Boerwinkle[31], Donald W. Bowden[32], Russell Bowler[33], Jennifer Brody[9], Ulrich Broeckel[34], Jai Broome[9], Karen Bunting[1], Esteban Burchard[35], Carlos Bustamante[36], Erin Buth[37], Brian Cade[38], Jonathan Cardwell[39], Vincent Carey[4], Cara Carty[40], Richard Casaburi[41], James Casella[12], Peter Castaldi[42], Mark Chaffin[3], Christy Chang[8], Yi-Cheng Chang[43], Daniel Chasman[44], Sameer Chavan[39], Bo-Juen Chen[1], Wei-Min Chen[45], Yii-Der Ida Chen[46], Michael Cho[4], Seung Hoan Choi[3], Lee-Ming Chuang[47], Mina Chung[48], Ren-Hua Chung[49], Clary Clish[3], Suzy Comhair[50], Matthew Conomos[37], Elaine Cornell[51], Adolfo Correa[52], Carolyn Crandall[41], James Crapo[53], L. Adrienne Cupples[54], Joanne Curran[55], Jeffrey Curtis[2], Brian Custer[56], Coleen Damcott[8], Dawood Darbar[57], Sayantan Das[2], Sean David[58], Colleen Davis[9], Michelle Daya[39], Mariza de Andrade[59], Lisa de las Fuentes[60], Michael DeBaun[61], Ranjan Deka[62], Dawn DeMeo[4], Scott Devine[8], Qing Duan[63], Ravi Duggirala[64], Jon Peter Durda[51], Susan Dutcher[65], Charles Eaton[66], Lynette Ekunwe[10], Adel El Boueiz[67], Patrick Ellinor[26], Leslie Emery[9], Serpil Erzurum[18], Charles Farber[45], Tasha Fingerlin[68], Matthew Flickinger[2], Myriam Fornage[31], Nora Franceschini[69], Chris Frazar[9], Mao Fu[8], Stephanie M. Fullerton[9], Lucinda Fulton[65], Stacey Gabriel[3], Weiniu Gan[24], Shanshan Gao[39], Yan Gao[10], Margery Gass[70], Bruce Gelb[71], Xiaoqi (Priscilla) Geng[2], Mark Geraci[72], Soren Germer[1], Robert Gerszten[73], Auyon Ghosh[4], Richard Gibbs[74], Chris Gignoux[16], Mark Gladwin[27], David Glahn[75], Stephanie Gogarten[9], Da-Wei Gong[8], Harald Goring[76], Sharon Graw[19], Daniel Grine[39], C. Charles Gu[65], Yue Guan[8], Xiuqing Guo[46], Namrata Gupta[3], Jeff Haessler[70], Michael Hall[10], Daniel Harris[8], Nicola L. Hawley[77], Jiang He[78], Ben Heavner[37], Susan Heckbert[9], Ryan Hernandez[35,79], David Herrington[80], Craig Hersh[81], Bertha Hidalgo[15], James Hixson[31], Brian Hobbs[4], John Hokanson[39], Elliott Hong[8], Karin Hoth[82], Chao (Agnes) Hsiung[83], Yi-Jen Hung[84], Haley Huston[85], Chii Min Hwu[86], Marguerite Ryan Irvin[15], Rebecca Jackson[87], Deepti Jain[9], Cashell Jaquish[24], Min A Jhun[2], Jill Johnsen[88], Andrew Johnson[24], Craig Johnson[9], Rich Johnston[7], Kimberly Jones[12], Hyun Min Kang[89], Robert Kaplan[90], Sharon Kardia[2], Sekar Kathiresan[3], Shannon Kelly[56], Eimear Kenny[71], Michael Kessler[8], Alyna Khan[9], Wonji Kim[91], Greg Kinney[39], Barbara Konkle[85], Charles Kooperberg[70], Holly Kramer[92], Christoph Lange[93], Ethan Lange[39], Leslie Lange[39], Cathy Laurie[9], Cecelia Laurie[9], Meryl LeBoff[4], Jiwon Lee[4], Seunggeun Shawn Lee[2], Wen-Jane Lee[86], Jonathon LeFaive[2], David Levine[9], Dan Levy[24], Joshua Lewis[8], Xiaohui Li[46], Yun Li[63], Henry Lin[46], Honghuang Lin[94], Keng Han Lin[2], Xihong Lin[95], Simin Liu[96], Yongmei Liu[97], Yu Liu[98], Ruth J.F. Loos[99], Steven Lubitz[26], Kathryn Lunetta[94], James Luo[24], Michael Mahaney[55], Barry Make[12], Ani Manichaikul[45], JoAnn Manson[4], Lauren Margolin[3], Lisa Martin[100], Susan Mathai[39], Rasika Mathias[12], Susanne May[37], Patrick McArdle[8], Merry-Lynn McDonald[15], Sean McFarland[91], Stephen McGarvey[66], Daniel McGoldrick[9], Caitlin McHugh[37], Hao Mei[10], Luisa Mestroni[19], Deborah A Meyers[101], Julie Mikulla[24], Nancy Min[10],

Mollie Minear[24], Ryan L Minster[27], Braxton D. Mitchell[8], Matt Moll[42], May E. Montasser[8], Courtney Montgomery[102], Arden Moscati[71], Solomon Musani[52], Stanford Mwasongwe[10], Josyf C Mychaleckyj[45], Girish Nadkarni[71], Rakhi Naik[12], Take Naseri[103], Pradeep Natarajan[3,26,91], Sergei Nekhai[104], Sarah C. Nelson[37], Bonnie Neltner[39], Deborah Nickerson[9], Kari North[63], Jeff O'Connell[8], Tim O'Connor[8], Heather Ochs-Balcom[105], David Paik[106], Nicholette Palmer[107], James Pankow[108], George Papanicolaou[24], Afshin Parsa[8], Juan Manuel Peralta[64], Marco Perez[16], James Perry[8], Ulrike Peters[109], Patricia Peyser[2], Lawrence S Phillips[7], Toni Pollin[8], Wendy Post[110], Julia Powers Becker[111], Meher Preethi Boorgula[39], Michael Preuss[71], Bruce Psaty[9], Pankaj Qasba[24], Dandi Qiao[4], Zhaohui Qin[7], Nicholas Rafaels[39], Laura Raffield[112], Vasan S. Ramachandran [94], D.C. Rao[65], Laura Rasmussen-Torvik[113], Aakrosh Ratan[45], Susan Redline[4], Robert Reed[8], Elizabeth Regan[53], Alex Reiner[109], Muagututi'a Sefuiva Reupena[114], Ken Rice[9], Stephen Rich[45], Dan Roden[115], Carolina Roselli[3], Jerome Rotter[46], Ingo Ruczinski[12], Pamela Russell[39], Sarah Ruuska[85], Kathleen Ryan[8], Ester Cerdeira Sabino[116], Danish Saleheen[20], Shabnam Salimi[8], Steven Salzberg[12], Kevin Sandow[117], Vijay G. Sankaran[3,118], Christopher Scheller[2], Ellen Schmidt[2], Karen Schwander[65], David Schwartz[39], Frank Sciurba[27], Christine Seidman[119], Jonathan Seidman[120], Vivien Sheehan[121], Stephanie L. Sherman[122], Amol Shetty[8], Aniket Shetty[39], Wayne Hui-Heng Sheu[86], M. Benjamin Shoemaker[123], Brian Silver[124], Edwin Silverman[4], Jennifer Smith[2], Josh Smith[9], Nicholas Smith[125], Tanja Smith[1], Sylvia Smoller[90], Beverly Snively[126], Michael Snyder[16], Tamar Sofer[4], Nona Sotoodehnia[9], Adrienne M. Stilp[9], Garrett Storm[39], Elizabeth Streeten[8], Jessica Lasky Su[4], Yun Ju Sung[65], Jody Sylvia[4], Adam Szpiro[9], Carole Sztalryd[8], Daniel Taliun[2], Hua Tang[127], Margaret Taub[12], Kent D. Taylor[128], Matthew Taylor[19], Simeon Taylor[8], Marilyn Telen[14], Timothy A. Thornton[9], Machiko Threlkeld[129], Lesley Tinker[70], David Tirschwell[9], Sarah Tishkoff[130], Hemant Tiwari[131], Catherine Tong[132], Russell Tracy[133], Michael Tsai[108], Dhananjay Vaidya[12], David Van Den Berg[134], Peter VandeHaar[2], Scott Vrieze[108,135], Tarik Walker[39], Robert Wallace[82], Avram Walts[39], Fei Fei Wang[9], Heming Wang[4], Karol Watson[41], Daniel E. Weeks[27], Bruce Weir[9], Scott Weiss[4], Lu-Chen Weng[26], Jennifer Wessel[136], Cristen Willer[137], Kayleen Williams[37], L. Keoki Williams[138], Carla Wilson[4], James Wilson[139], Quenna Wong[9], Joseph Wu[106], Huichun Xu[8], Lisa Yanek[12], Ivana Yang[39], Rongze Yang[8], Norann Zaghloul[8], Maryam Zekavat[3], Yingze Zhang[140], Snow Xueyan Zhao[53], Wei Zhao[141], Degui Zhi[31], Xiang Zhou[2], Xiaofeng Zhu[142], Michael Zody[1], Sebastian Zoellner[2]

1 - New York Genome Center, New York, New York, US; 2 - University of Michigan, Ann Arbor, Michigan, US; 3 - Broad Institute, Cambridge, Massachusetts, US; 4 - Brigham & Women's Hospital, Boston, Massachusetts, US; 5 - Cedars Sinai, Los Angeles, California, US; 6 - Children's Hospital of Philadelphia, University of Pennsylvania, Philadelphia, Pennsylvania, US; 7 - Emory University, Atlanta, Georgia, US; 8 - University of Maryland, Baltimore, Maryland, US; 9 - University of Washington, Seattle, Washington, US; 10 - University of Mississippi, Jackson, Mississippi, US; 11 - National Institutes of Health, Bethesda, Maryland, US; 12 - Johns Hopkins University, Baltimore, Maryland, US; 13 - University of Kentucky, Lexington, Kentucky, US; 14 - Duke University, Durham, North Carolina, US; 15 - University of Alabama, Birmingham, Alabama, US; 16 - Stanford University, Stanford, California, US; 17 - University of Wisconsin Milwaukee, Milwaukee, Wisconsin, US; 18 - Cleveland Clinic, Cleveland, Ohio, US; 19 - University of Colorado Anschutz Medical Campus, Aurora, Colorado, US; 20 - Columbia

University, New York, New York, US; 21 - The Emmes Corporation, LTRC, Rockville, Maryland, US; 22 - Cleveland Clinic, Quantitative Health Sciences, Cleveland, Ohio, US; 23 - Johns Hopkins University, Medicine, Baltimore, Maryland, US; 24 - National Heart, Lung, and Blood Institute, National Institutes of Health, Bethesda, Maryland, US; 25 - Boston University, Massachusetts General Hospital, Boston University School of Medicine, Boston, Massachusetts, US; 26 - Massachusetts General Hospital, Boston, Massachusetts, US; 27 - University of Pittsburgh, Pittsburgh, Pennsylvania, US; 28 - Fundação de Hematologia e Hemoterapia de Pernambuco - Hemope, Recife, BR; 29 - University of Washington, Cardiovascular Health Research Unit, Department of Medicine, Seattle, Washington, US; 30 - University of Texas Rio Grande Valley School of Medicine, Human Genetics, Brownsville, Texas, US; 31 - University of Texas Health at Houston, Houston, Texas, US; 32 - Wake Forest Baptist Health, Department of Biochemistry, Winston-Salem, North Carolina, US; 33 - National Jewish Health, National Jewish Health, Denver, Colorado, US; 34 - Medical College of Wisconsin, Milwaukee, Wisconsin, US; 35 - University of California, San Francisco, San Francisco, California, US; 36 - Stanford University, Biomedical Data Science, Stanford, California, US; 37 - University of Washington, Biostatistics, Seattle, Washington, US; 38 - Brigham & Women's Hospital, Brigham and Women's Hospital, Boston, Massachusetts, US; 39 - University of Colorado at Denver, Denver, Colorado, US; 40 - Washington State University, Pullman, Washington, US; 41 - University of California, Los Angeles, Los Angeles, California, US; 42 - Brigham & Women's Hospital, Medicine, Boston, Massachusetts, US; 43 - National Taiwan University, Taipei, TW; 44 - Brigham & Women's Hospital, Division of Preventive Medicine, Boston, Massachusetts, US; 45 - University of Virginia, Charlottesville, Virginia, US; 46 - Lundquist Institute, Torrance, California, US; 47 - National Taiwan University, National Taiwan University Hospital, Taipei, TW; 48 - Cleveland Clinic, Cleveland Clinic, Cleveland, Ohio, US; 49 - National Health Research Institute Taiwan, Miaoli County, TW; 50 - Cleveland Clinic, Immunity and Immunology, Cleveland, Ohio, US; 51 - University of Vermont, Burlington, Vermont, US; 52 - University of Mississippi, Medicine, Jackson, Mississippi, US; 53 - National Jewish Health, Denver, Colorado, US; 54 - Boston University, Biostatistics, Boston, Massachusetts, US; 55 - University of Texas Rio Grande Valley School of Medicine, Brownsville, Texas, US; 56 - Vitalant Research Institute, San Francisco, California, US; 57 - University of Illinois at Chicago, Chicago, Illinois, US; 58 - University of Chicago, Chicago, Illinois, US; 59 - Mayo Clinic, Health Sciences Research, Rochester, Minnesota, US; 60 - Washington University in St Louis, Department of Medicine, Cardiovascular Division, St. Louis, Missouri, US; 61 - Vanderbilt University, Nashville, Tennessee, US; 62 - University of Cincinnati, Cincinnati, Ohio, US; 63 - University of North Carolina, Chapel Hill, North Carolina, US; 64 - University of Texas Rio Grande Valley School of Medicine, Edinburg, Texas, US; 65 - Washington University in St Louis, St Louis, Missouri, US; 66 - Brown University, Providence, Rhode Island, US; 67 - Harvard University, Channing Division of Network Medicine, Cambridge, Massachusetts, US; 68 - National Jewish Health, Center for Genes, Environment and Health, Denver, Colorado, US; 69 - University of North Carolina, Epidemiology, Chapel Hill, North Carolina, US; 70 - Fred Hutchinson Cancer Research Center, Seattle, Washington, US; 71 - Icahn School of Medicine at Mount Sinai, New York, New York, US; 72 - Indiana University, Medicine, Indianapolis, Indiana, US; 73 - Beth Israel Deaconess Medical Center, Boston, Massachusetts, US; 74 - Baylor College of Medicine Human Genome Sequencing Center,

Houston, Texas, US; 75 - Yale University, New Haven, Connecticut, US; 76 - University of Texas Rio Grande Valley School of Medicine, San Antonio, Texas, US; 77 - Yale University, Department of Chronic Disease Epidemiology, New Haven, Connecticut, US; 78 - Tulane University, New Orleans, Louisiana, US; 79 - McGill University, Montreal, CA; 80 - Wake Forest Baptist Health, Winston-Salem, North Carolina, US; 81 - Brigham & Women's Hospital, Channing Division of Network Medicine, Boston, Massachusetts, US; 82 - University of Iowa, Iowa City, Iowa, US; 83 - National Health Research Institute Taiwan, Institute of Population Health Sciences, NHRI, Miaoli County, TW; 84 - Tri-Service General Hospital National Defense Medical Center, TW; 85 - Blood Works Northwest, Seattle, Washington, US; 86 - Taichung Veterans General Hospital Taiwan, Taichung City, TW; 87 - Ohio State University Wexner Medical Center, Internal Medicine, DIvision of Endocrinology, Diabetes and Metabolism, Columbus, Ohio, US; 88 - Blood Works Northwest, University of Washington, Seattle, Washington, US; 89 - University of Michigan, Biostatistics, Ann Arbor, Michigan, US; 90 - Albert Einstein College of Medicine, New York, New York, US; 91 - Harvard University, Cambridge, Massachusetts, US; 92 - Loyola University, Public Health Sciences, Maywood, Illinois, US; 93 - Harvard School of Public Health, Biostats, Boston, Massachusetts, US; 94 - Boston University, Boston, Massachusetts, US; 95 - Harvard School of Public Health, Boston, Massachusetts, US; 96 - Brown University, Epidemiology and Medicine, Providence, Rhode Island, US; 97 - Duke University, Cardiology, Durham, North Carolina, US; 98 - Stanford University, Cardiovascular Institute, Stanford, California, US; 99 - Icahn School of Medicine at Mount Sinai, The Charles Bronfman Institute for Personalized Medicine, New York, New York, US; 100 - George Washington University, Washington, District of Columbia, US; 101 - University of Arizona, Tucson, Arizona, US; 102 - Oklahoma Medical Research Foundation, Genes and Human Disease, Oklahoma City, Oklahoma, US; 103 - Ministry of Health, Government of Samoa, Apia, WS; 104 - Howard University, Washington, District of Columbia, US; 105 - University at Buffalo, Buffalo, New York, US; 106 - Stanford University, Stanford Cardiovascular Institute, Stanford, California, US; 107 - Wake Forest Baptist Health, Biochemistry, Winston-Salem, North Carolina, US; 108 - University of Minnesota, Minneapolis, Minnesota, US; 109 - Fred Hutchinson Cancer Research Center, University of Washington, Seattle, Washington, US; 110 - Johns Hopkins University, Cardiology/Medicine, Baltimore, Maryland, US; 111 - University of Colorado at Denver, Medicine, Denver, Colorado, US; 112 - University of North Carolina, Genetics, Chapel Hill, North Carolina, US; 113 - Northwestern University, Chicago, Illinois, US; 114 - Lutia I Puava Ae Mapu I Fagalele, Apia, WS; 115 - Vanderbilt University, Medicine, Pharmacology, Biomedicla Informatics, Nashville, Tennessee, US; 116 - Universidade de Sao Paulo, Faculdade de Medicina, Sao Paulo, BR; 117 - Lundquist Institute, TGPS, Torrance, California, US; 118 - Harvard University, Division of Hematology/Oncology, Cambridge, Massachusetts, US; 119 - Harvard Medical School, Genetics, Boston, Massachusetts, US; 120 - Harvard Medical School, Boston, Massachusetts, US; 121 - Baylor College of Medicine, Pediatrics, Houston, Texas, US; 122 - Emory University, Human Genetics, Atlanta, Georgia, US; 123 - Vanderbilt University, Medicine/Cardiology, Nashville, Tennessee, US; 124 - UMass Memorial Medical Center, Worcester, Massachusetts, US; 125 - University of Washington, Epidemiology, Seattle, Washington, US; 126 - Wake Forest Baptist Health, Biostatistical Sciences, Winston-Salem, North Carolina, US; 127 - Stanford University, Genetics, Stanford, California, US; 128 - Lundquist Institute, Institute for Translational Genomics and Populations Sciences, Torrance,

California, US; 129 - University of Washington, University of Washington, Department of Genome Sciences, Seattle, Washington, US; 130 - University of Pennsylvania, Genetics, Philadelphia, Pennsylvania, US; 131 - University of Alabama, Biostatistics, Birmingham, Alabama, US; 132 - University of Washington, Department of Biostatistics, Seattle, Washington, US; 133 - University of Vermont, Pathology & Laboratory Medicine, Burlington, Vermont, US; 134 - University of Southern California, USC Methylation Characterization Center, University of Southern California, California, US; 135 - University of Colorado at Boulder, Boulder, Colorado, US; 136 - Indiana University, Epidemiology, Indianapolis, Indiana, US; 137 - University of Michigan, Internal Medicine, Ann Arbor, Michigan, US; 138 - Henry Ford Health System, Detroit, Michigan, US; 139 - Beth Israel Deaconess Medical Center, Cardiology, Boston, Massachusetts, US; 140 - University of Pittsburgh, Medicine, Pittsburgh, Pennsylvania, US; 141 - University of Michigan, Department of Epidemiology, Ann Arbor, Michigan, US; 142 - Case Western Reserve University, Department of Population and Quantitative Health Sciences, Cleveland, Ohio, US
